# Population differences in wearable device wear time: Rescuing data to address biases and advance health equity

**DOI:** 10.64898/2026.03.06.26347799

**Authors:** Eric Hurwitz, Evan Connelly, Miriam Sklerov, Hiral Master, Harry Hochheiser, Zachary Butzin-Dozier, Jessilyn Dunn, Melissa A. Haendel

## Abstract

Wearable devices present transformative opportunities for personalized healthcare through continuous monitoring of digital biomarkers; however, individual variations in device wear time could mask or otherwise impact signal identification. Despite the widespread adoption of wearable devices in research, no comprehensive framework exists for understanding how wear time varies across populations or for addressing wear time-related biases in analysis. Using Fitbit data from 11,901 participants in the *All of Us* Research Program, we conducted the first large-scale systematic assessment of wearable device wear time across demographics, social determinants of health, lifestyle factors, mental health symptoms, and disease. Our findings revealed that wear time was higher among males and increased with age, income, and education, but decreased with depressive, anxiety, and anhedonia symptoms, with reductions more pronounced following clinical diagnoses compared to symptom-based classifications. Individuals with chronic conditions displayed differential levels of wear time compared to healthy controls. Critically, we demonstrate that the widely used ≥10-hour daily compliance threshold, while appropriate for some research contexts, can disproportionately exclude days of data from disease populations: among individuals with major depressive disorder, 74.4% of data days were excluded compared to 20.9% for controls. We propose a flexible methodological framework including standard compliance thresholds, wear time covariate adjustment, metric normalization, propensity score matching, and adaptive thresholds that can be applied individually or in combination to optimize wearable data retention across diverse research contexts. These findings establish wear time as a critical methodological consideration for wearable device research and provide guidance for advancing equitable and rigorous digital health analytics.

## Introduction

The rise of wearable device ownership, such as Fitbits, has led to significant advancements for personal health^1^. Because wearables monitor the same individual in a continuous and longitudinal manner, their use is particularly exciting in the movement towards personalized medicine^2^. Wearable devices collect digital biomarkers to capture data passively and objectively in non-clinical settings. Digital biomarkers from wearables present an opportunity to transform healthcare and research capabilities with low-cost and convenient technology to provide personalized insights about health^3^.

Prior large-scale longitudinal, observational studies have shown physical activity^4^ and sleep patterns^5^ are associated with incident chronic disease. Additionally, machine learning using digital biomarkers from wearables have clinical applications ranging from acute event detection (e.g., infection^6–8^) to management of chronic disease (e.g., cardiovascular disease^9,10^ and diabetes^11,12^). While these studies established a relationship between sensor-derived digital biomarkers (e.g., heart rate, physical activity, and sleep) and disease through statistical and machine learning methods, how to methodologically account for individual variations in device wear time, which could mask or otherwise impact signal identification, remains unresolved. Prior studies looking at wearable device wear time have mainly taken place in general, disease-agnostic populations, and focus on non-biomedical contexts to study user behavior with technology^13–18^. It is crucial to investigate wearable device wear time across participants with a spectrum of health and disease states to maximize the clinical utility of wearable devices.

Applications of wearable devices and their data are clinical trials, observational studies, and population health research, where understanding wear time patterns is essential for maximizing the return on data collection investments. To maximize the utility of these data, it is necessary to consider not only sensor-derived biomarkers (e.g., heart rate, physical activity, and sleep) but also wear time itself, which may serve as an indicator of clinical status and outcomes^19^. Currently, digital health studies typically filter on ‘compliant’ data days (≥10 hours wear time), which discards data that can still contribute valid signal when accounted for and creates sampling bias against groups with lower device adherence, while also incurring financial costs from data exclusion. The ≥10-hour threshold originates from the NHANES accelerometry literature, where it was established to ensure sufficient data capture for reliable estimation of daily activity patterns in population surveillance studies. While this criterion remains appropriate for certain applications, particularly when capturing a representative single-day snapshot of behavior in healthy populations, its universal application to disease-specific research may inadvertently discard clinically meaningful data^18^. In clinical contexts, reduced wear time itself may reflect disease burden, symptom severity, or treatment effects, making these observations particularly valuable rather than expendable. For example, a study including wearable devices in breast cancer patients undergoing chemotherapy suggested that drug toxicity may contribute to decreased engagement with the device^20^. Additionally, an observational study using wearable devices, the electronic Framingham Heart Study, found that individuals with depressive symptoms were likely to have lower wear time of their device^21^. Overall, levels of wearable device wear time is likely non-random and can offer valuable insights in the context of study participants during data collection.

This study aimed to systematically characterize how wearable device wear time varies by demographics, social determinants of health (SDoH), lifestyle factors, mental health symptoms, and a range of chronic disease categories to establish wear time as a critical methodological consideration for wearable device research. We propose a flexible analytical framework for maximizing wearable device data retention, including adjusting for hours of daily wear time as a covariate alongside complementary approaches such as metric normalization, propensity score matching, and adaptive wear time thresholds. We leveraged Fitbit, survey, and electronic health record (EHR) data from the *All of Us* (AoU) research program, an initiative that aims to gather multiple health-related streams of information in one million Americans with a focus on patient populations that are under-represented in research^22^. This comprehensive approach provides both a foundational characterization of wear time patterns across diverse populations and practical methodological guidance for optimizing wearable device research while advancing health equity.

## Methods

### Data source and platform

Data described in this study were analyzed using the AoU Controlled Tier v7 dataset. All analyses were conducted in the Researcher Workbench cloud platform using R. Fitbit data in AoU v7 is collected under a bring-your-own-device model, where study participants who already own a Fitbit share their data by linking their device to the AoU portal.

### Ethical considerations

The protocol for the AoU study was reviewed by the institutional review board of AoU (protocol 2021-02-TN-001). The institutional review board follows the regulations and guidance of the National Institutes of Health Office for Human Research Protections for all studies, ensuring that the rights and welfare of research participants are overseen and protected uniformly. The informed consent process states that participants have the option to withdraw at any time. Privacy of participant data is maintained in the following three ways: 1) storing data on protected computers, 2) preventing researchers from seeing identifiable patient information, such as name or social security number, 3) having researchers sign a contract they won’t try to identify participants. Furthermore, access to the AoU dataset is only available through the Researcher Workbench, which is only available to researchers who have completed the requisite training at institutions with a signed Data Use Agreement. For compensation, participants are offered $25 one-time in the form of cash, gift card, or an electronic voucher if they are asked and decide to go to an AoU partner center for physical measurements to give blood, saliva, or urine samples.

### Statistical analyses

We assessed Fitbit wear time using two key research questions: 1) “How does the percentage of days individuals wear their Fitbit differ between groups X and Y?” and 2) “What is the difference in the daily hours of Fitbit wear time between groups X and Y?”. Analysis was performed in the same manner as our prior work^23^ and is described below.

For question 1, we calculated the percentage of days each individual wore their Fitbit using the number of days an individual had any recorded step data divided by the total number of days they could have worn their Fitbit (i.e., the difference in days between the first and last day each person had any record of Fitbit data). Analysis of Fitbit wear time was conducted using either linear regression or linear mixed-effects models using the *lmer* package in R^24^. Linear mixed-effects models were used in the case where there was more than one measurement per person to avoid the issue of correlation between data points. Please refer to Supplemental Methods in the *Data preprocessing for covariates section* for details about how covariates were obtained in AoU. For question 2, we determined the daily Fitbit wear time (in hours) per individual by counting the number of hours in a day with non-zero step counts, consistent with prior work^4,25^. To optimize computational efficiency in the Researcher Workbench, we computed mean daily wear hours per participant across relevant time windows for each analysis and applied linear regression models. Consistent with our earlier approach, we used linear mixed-effects models to account for within-participant correlation when multiple observations were present. Statistical significance was assessed at α=0.05 with Bonferroni correction applied to account for multiple comparisons within each analysis (e.g., when comparing across 6 racial/ethnic groups, the corrected significance threshold was 0.05/5=0.01). All models from both questions 1 and 2 were assessed at a significance level of 0.05 with Bonferroni correction^26^.

### Data preprocessing and statistical analysis comparing Fitbit wear time by demographics, SDoH, and lifestyle factors

Age, sex, and race/ethnicity were obtained for each individual using the Person table in AoU. For each individual with any Fitbit data available in the AoU, we calculated 1) the percentage of days and 2) the number of daily hours they wore their Fitbit as described above. For the two calculations, we filtered on individuals with at least 30 total days of Fitbit step data (since step data was used to estimate wear time) to ensure a reliable pattern of wear time^6,7^. For instance, if someone had only 2 days of available Fitbit data and wore their Fitbit both days, they would show 100% wear time which would be misleading due to the limited data. When preparing data for the number of daily hours each individual wore their Fitbit, we calculated the average hours of wear time per person across time periods of interest and then performed linear regression models. Race/ethnicity was determined by merging the race and ethnicity columns in the Person table. Furthermore, we recategorized individuals’ race/ethnicity groups with less than 20 participants as *Other NH* race/ethnicity in order to comply with AoU’s requirement of at least 20 participants per group to mitigate the risk of participant reidentification^27^.

### Preparing and analyzing mood, anxiety, and anhedonia symptom survey data

Survey questions about individual levels of mood, anxiety, and anhedonia symptoms were assessed using all six versions deployed from May 2020 through February 2021 of the *COVID-19 Participant Experience (COPE)* survey^28^. Please refer to Supplemental Methods in the *Preprocessing depressive, anxiety, and anhedonia symptoms* for details about how data were obtained and prepared in AoU.

### Preparing and analyzing EHR data amongst those with major depressive disorder (MDD) and anxiety disorder (AD)

Individuals were identified as part of the MDD cohort following the Depression phenotype workspace available in AoU^29^. Please refer to Supplemental Methods in the *Preprocessing major depressive disorder and anxiety disorder diagnosis data* for details about how data were obtained and prepared in AoU.

### Preprocessing and analysis of individuals with mental disorders

Individuals were classified as having a mental disorder if they had any EHR diagnosis of at least one mental disorder listed in Table S1 based on the presence of an EHR diagnosis using the Conditions table in AoU. Groups of mental disorders included were selected based on the Phemap and Phecodes as described previously^4,5,30^. Specific mental disorders were excluded from analysis if there were less than 20 individuals to stay in compliance with the AoU dissemination policy^27^. For each individual, we first identified the date of diagnosis for each mental disorder and then determined the percentage of days their Fitbit was worn between the date of diagnosis for that mental disorder and the last recorded day of any Fitbit data in AoU. To ensure that the data analyzed was during when individuals had the mental disorder, Fitbit data was filtered after the index date for each mental disorder. For individuals diagnosed with mental disorders before Fitbit devices became commercially available in 2009, we reset the index date to January 1, 2010. A cohort of individuals without mental disorders was created by excluding those who did not have a diagnosis of any mental disorder listed in Table S1, where the index date for these individuals was the median number of days each disorder was diagnosed after the first day of Fitbit data in the cohort with mental disorders, consistent with our prior work^25^. To temporally align comparisons between individuals with and without mental disorders, we calculated the median index date among individuals with mental disorders (July 19, 2016) and assigned this date as the index date for control individuals without mental disorders. We also filtered on individuals with at least 30 total days of Fitbit data (in total and with their mental disorder) to ensure a reliable pattern of wear time. We then performed a linear mixed-effects model with person ID as the random effect (since some individuals could have more than one mental disorder) to compare the 1) percentage of days and 2) hours per day the Fitbit was worn between those without a mental disorder and individuals with each mental disorder at a significance level of 0.05 with Bonferroni adjustment and adjusted for age, sex, race/ethnicity, annual income, education level, and health insurance status.

To compare differences in the percentage of Fitbit wear time before and after the diagnosis of each mental disorder, we determined the possible number of days each individual could have worn their Fitbit between the first recorded day of any Fitbit data in AoU and the date of mental disorder diagnosis, counted the number of days each individual had any step data between those two dates, and calculated the percentage of days each individual wore their Fitbit before diagnosis. This process was repeated in a similar manner to determine the percentage of days each individual wore their Fitbit after being diagnosed with a mental disorder, anchoring between the date of diagnosis and the last date of any recorded Fitbit data for that person in AoU. A similar process was conducted to determine the number of hours per day. The same methods were performed for each mental disorder. Linear mixed-effects models with person ID as a random effect tested the interaction between mental disorder diagnosis and time period (before/after index date) on Fitbit wear time 1) percentage and 2) hours per day, adjusting for age, sex, race/ethnicity, annual income, education level, and health insurance status with Bonferroni correction.

### Preprocessing and analysis of individuals with different types of disorders

Individuals were identified as having one or multiple of the chronic disorders listed in Table S2 based on the presence of an EHR diagnosis using the conditions table in AoU. All groups of disorders included were selected based on the Phemap and Phecodes^4,5,30^. Disorders with counts of less than 20 individuals were excluded due to AoU privacy policies^27^. We also excluded *Congenital anomalies* because these are present from birth, making it impossible to compare Fitbit wear time before versus after diagnosis. We followed the same steps as described with mental disorders to determine the percentage of days and hours per day each individual wore their Fitbit with each disorder and compared that to a healthy control cohort, defined as someone who was not diagnosed with any of the individuals without any of the conditions listed in Table S2. The analysis was done using a linear mixed-effects model with person ID as the random effect adjusted for age, sex, race/ethnicity, annual income, education level, and health insurance status at a significance level of 0.05. We also assessed differences in wear time before and after the diagnosis of each condition using an interaction model as described above with Bonferroni correction.

### Wear time adjustment methods

Variability in device wear time across individuals and clinical populations presents a methodological challenge for wearable device research, as standard compliance filtering may systematically exclude data from populations with reduced wear adherence. To address this, we evaluated five analytical approaches for handling wear time variability, each with distinct assumptions, strengths, and limitations (Table 1). These methods are not mutually exclusive and can be selected or combined based on the research question, outcome of interest, and study population. Previous evaluations of observational health data have addressed similar concerns regarding heterogeneous monitoring^31–34^.

**Table 1.** Methodological approaches for handling wear time in wearable device data analysis.

| Method | Description | Pros | Cons | When to choose |
| --- | --- | --- | --- | --- |
| **Standard compliance filtering (e.g., $\geq 10$ hrs/day)** | Excludes person-days with fewer than 10 hours of device wear time, retaining only days meeting a minimum wear duration for analysis. | Widely established; enables cross-study comparability<br>Ensures sufficient data per day for reliable metric estimation<br>Simple to implement and interpret | Disproportionately excludes data from specific populations<br>Introduces systematic bias through data exclusion<br>Reduced statistical power and smaller analytic samples | When cross-study comparability is required<br>When estimating representative daily activity patterns in generally healthy populations |
| **Covariate adjustment** | Includes all person-days regardless of wear duration and adds daily wear time (in hours) as a covariate in statistical models to control for differential wear across groups. | Retains 100% of participant-days regardless of wear duration<br>Statistically controls for wear time differences across groups<br>Maximized statistical power and sample representativeness<br>Straightforward to implement in standard regression frameworks | Assumes a linear (or specified) relationship between wear time and outcomes<br>May not fully address confounding if wear time-outcome relationship is complex<br>Absolute activity estimates still influenced by wear duration<br>Requires sufficient variation in wear time for stable estimation | When maximizing statistical power and sample representativeness is the priority, particularly for group-level comparisons where data loss would disproportionately affect one group (e.g., disease vs. control comparisons) |
| **Metric normalization** | Converts cumulative daily metrics into rate-based measures by dividing by hours of wear time (e.g., steps per hour rather than total daily steps), standardizing activity estimates across variable wear durations. | Directly adjusts metrics for variable wear duration<br>Provides intuitive, rate-based measures comparable across participants<br>Retains majority of data<br>Eliminates systematic inflation/deflation of cumulative metrics | Requires minimum wear time floor to avoid unstable rates from very short wear periods<br>Assumes activity rate is constant throughout the day<br>Some metrics (e.g., heart rate) may not be meaningfully normalized by wear time<br>Still excludes days with zero or near-zero wear time | When the outcome of interest scales directly with wear duration (e.g., step counts, active minutes) and the goal is an intuitive, rate-based measure comparable across participants with variable wear durations |
| **Propensity matching** | Matches participants or person-days across comparison groups on wear time distributions using propensity scores, ensuring equivalent device exposure between groups prior to analysis. | Ensures comparison groups have equivalent wear time distributions<br>Creates balanced analytic samples, reducing confounding by wear time<br>Enables causal inference framing with well-matched cohorts<br>Particularly useful for case-control or exposure-outcome designs | Substantially reduces sample size<br>Excludes unmatched participants limiting generalizability<br>Matching quality depends on propensity model specification<br>Computationally intensive for large datasets | When the study design requires equivalent device exposure between comparison groups prior to analysis, such as in case-control or exposure-outcome studies where confounding by wear time must be eliminated rather than statistically adjusted |
| **Adaptive wear time threshold** | Applies a lower wear time threshold (e.g., 6 or 8 hours) calibrated so that the variable of interest does not differ significantly between comparison groups, balancing data retention with analytical validity. | ● Retains more data than standard $\geq 10$ hour threshold<br>● Threshold can be calibrated so key variables do not significantly differ between groups<br>● Balances data quality with data retention<br>● Flexible and study-specific; adaptable to different populations and outcomes | ● No universal consensus on optimal lower threshold<br>● Threshold selection may appear post-hoc without pre-registration<br>● Lower thresholds may reduce reliability of daily activity estimates<br>● Requires sensitivity analyses to demonstrate robustness across thresholds | ● When researchers seek to retain more data than the standard $\geq 10$ hours per day threshold while still ensuring minimum data quality per day, particularly when wear time differences between groups are expected to diminish at lower thresholds |

To demonstrate how each approach affects estimation of a well-established clinical relationship, we compared daily Fitbit step counts between individuals with and without MDD, a condition selected because reduced physical activity is a well-documented feature of depression. All models were fit as linear mixed-effects models with a random intercept for person ID and adjusted for age, race/ethnicity, and sex at birth. The five approaches were implemented as follows. First, standard compliance filtering retained only person-days meeting established quality criteria (≥10 hours of wear time, >100 steps, and <45,000 steps) and modeled total daily steps as the outcome. Second, covariate adjustment retained all person-days regardless of wear duration and included hours of daily wear time as an additional covariate, with total daily steps as the outcome. Third, metric normalization retained all person-days with non-zero wear time and used steps per hour of wear time as the outcome, directly standardizing activity by wear duration rather than adjusting for it statistically. Fourth, propensity score matching used nearest-neighbor 1:1 matching without replacement on hours of wear time at the person-day level to create comparison groups with equivalent wear time distributions, then modeled total daily steps on the matched sample. Fifth, adaptive threshold analysis systematically evaluated minimum wear time thresholds from 1 to 10 hours, at each threshold filtering the data accordingly and testing whether wear time remained significantly different between groups using Wilcoxon rank-sum tests, providing a sensitivity analysis of how threshold selection affects both data retention and residual confounding by wear time. Rather than selecting a single optimal threshold, this approach illustrates how researchers could calibrate a study-specific threshold at which wear time no longer differs significantly between comparison groups, though in the present MDD example, wear time differences persisted across all thresholds tested.

### Creating graphs

All graphs were created using the *ggplot2* package^35^ in R.

### Large Language models (LLMs)

ChatGPT (GPT-4o and GPT-3.5), developed by OpenAI^36^, and Claude (Sonnet 4.5, Opus 4.5), developed by Anthropic^37^, were used to edit some portions of the manuscript, including grammar, language, and synonyms. All recommendations from LLMs were reviewed by the author and were not used for the purpose of generating ideas or content.

## Results

### AoU cohort characteristics

Our AoU analysis included 11,901 individuals with available Fitbit and survey data (Figure 1). Of those, the majority of individuals were female (n=8,368, 70.3%). By age and race/ethnicity, the largest subgroups in our cohort were between 60-70 years old (n=2,666, 22.4%) and White non-Hispanic (NH) (n=9,665, 81.2%; Table 2). When examining SDoH, the majority of those in our cohort had an annual income between $100-150K (n=2,485, 20.9%), held an advanced degree as their highest level of education (n=4,634, 38.9%), and had health insurance coverage (n=11,602, 97.5%; Table 2). In contrast, the smallest groups included those who skipped the annual income survey question (n=224, 1.9%), skipped the highest education level survey question (n=160, 1.3%), and those without health insurance (n=148, 1.2%; Table 2). In regards to lifestyle, the majority of individuals have not smoked more than 100 cigarettes (n=7,369, 65.8%) and have consumed at least one alcoholic beverage (n=10,694, 95.4%) in their lifetime (Table 2).

**Figure 1:**
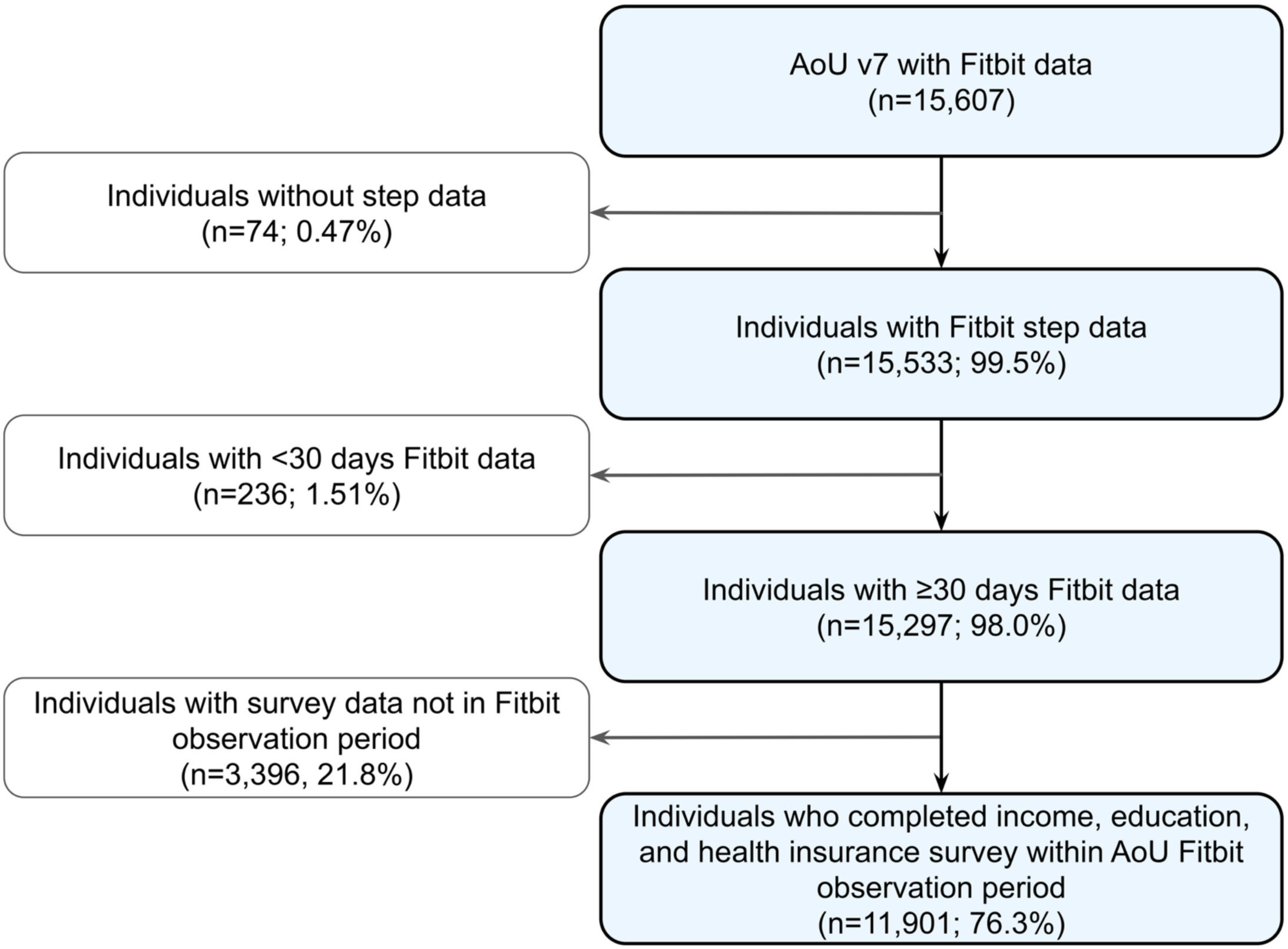
Flow diagram for our analytic cohort. AoU=All of Us *A flowchart describing the inclusion and exclusion criteria of our AoU v7 analytic cohort using Fitbit intra-day steps data*.

**Table 2:**
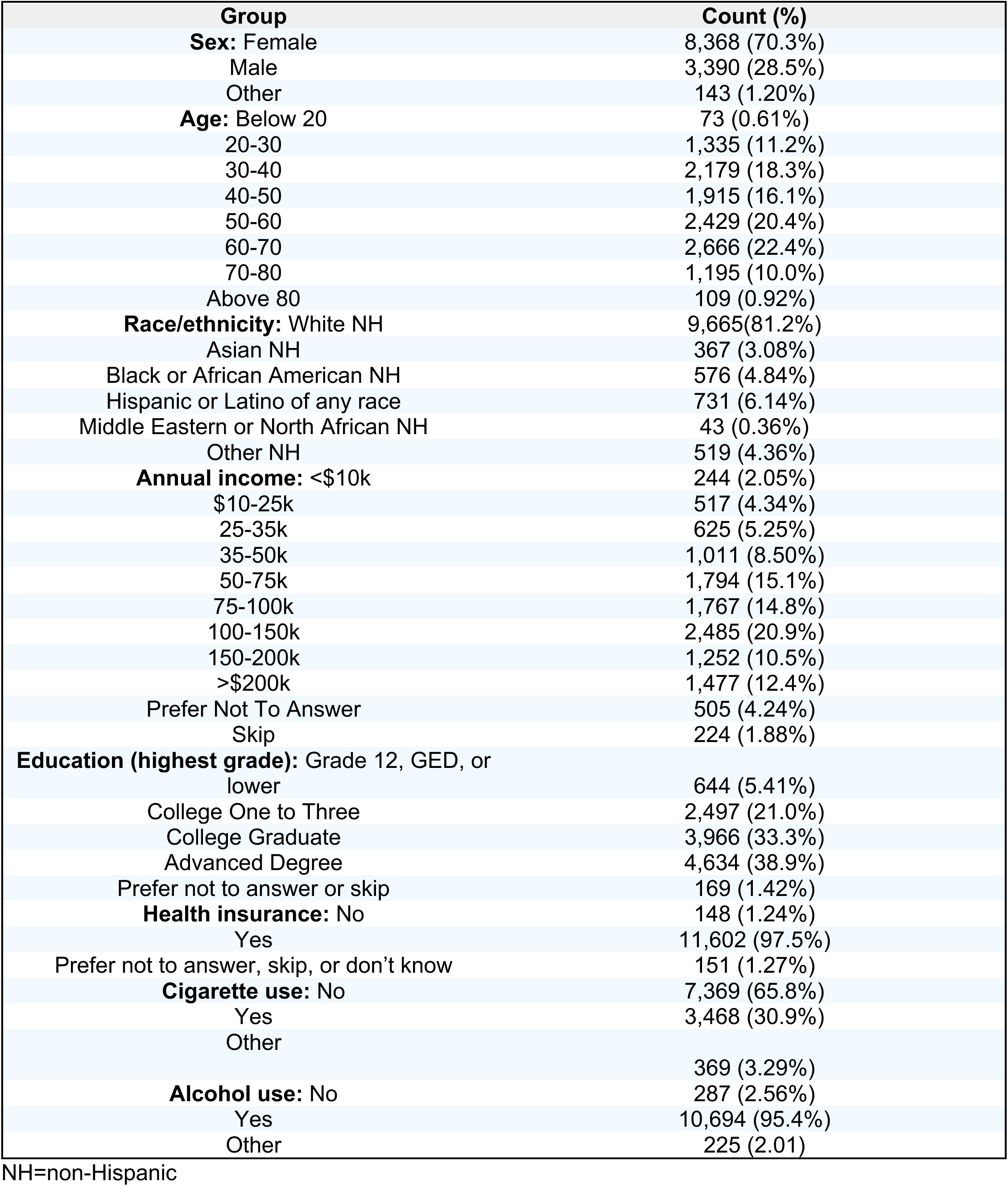
Descriptive statistics of the AoU cohort that contributed wearable device data.

### Wearable device wear time varies by demographics and social determinants of health

We first aimed to assess differences in Fitbit device wear time across the entire AoU cohort by demographics (sex, age, race/ethnicity), SDoH (income, education, health insurance status), and lifestyle factors (cigarette and alcohol use). This analysis was conducted by comparing the percentage of days the Fitbit was worn across different groups. The results showed a significant difference in the percentage of days Fitbits were worn with increasing wearable wear time with increasing age and the highest percentage of wear time among individuals aged 60-70 years old (Figure 2A). Furthermore, males exhibited increased wear time relative to females (estimate=2.83%, 95% confidence interval [CI]=1.82, 3.84%, *P*<0.001) while those who were considered other sex at birth did not exhibit different wear time relative to females (estimate=0.17%, 95% CI=-4.03, 4.36%, *P*>0.05) (Figure 2B). Individuals who were Hispanic/Latino of any race displayed decreased wear time compared to White NH counterparts (estimate=-6.19%, 95% CI=-8.10, –4.28%, *P*<0.001; Figure 2C). Regarding income and education, individuals with higher income and higher education had increased percentages of wear time (Figures 2D and 2E). We also detected that those with health insurance wear their device significantly more than those without health insurance (estimate=6.54%, 95% CI=2.42, 10.7%, *P*<0.01; Figure 2F). When assessing lifestyle differences, the percentage of wear time did not differ significantly between individuals who smoked or consumed alcohol and those who did not. (Figures S1A and S1B).

**Figure 2:**
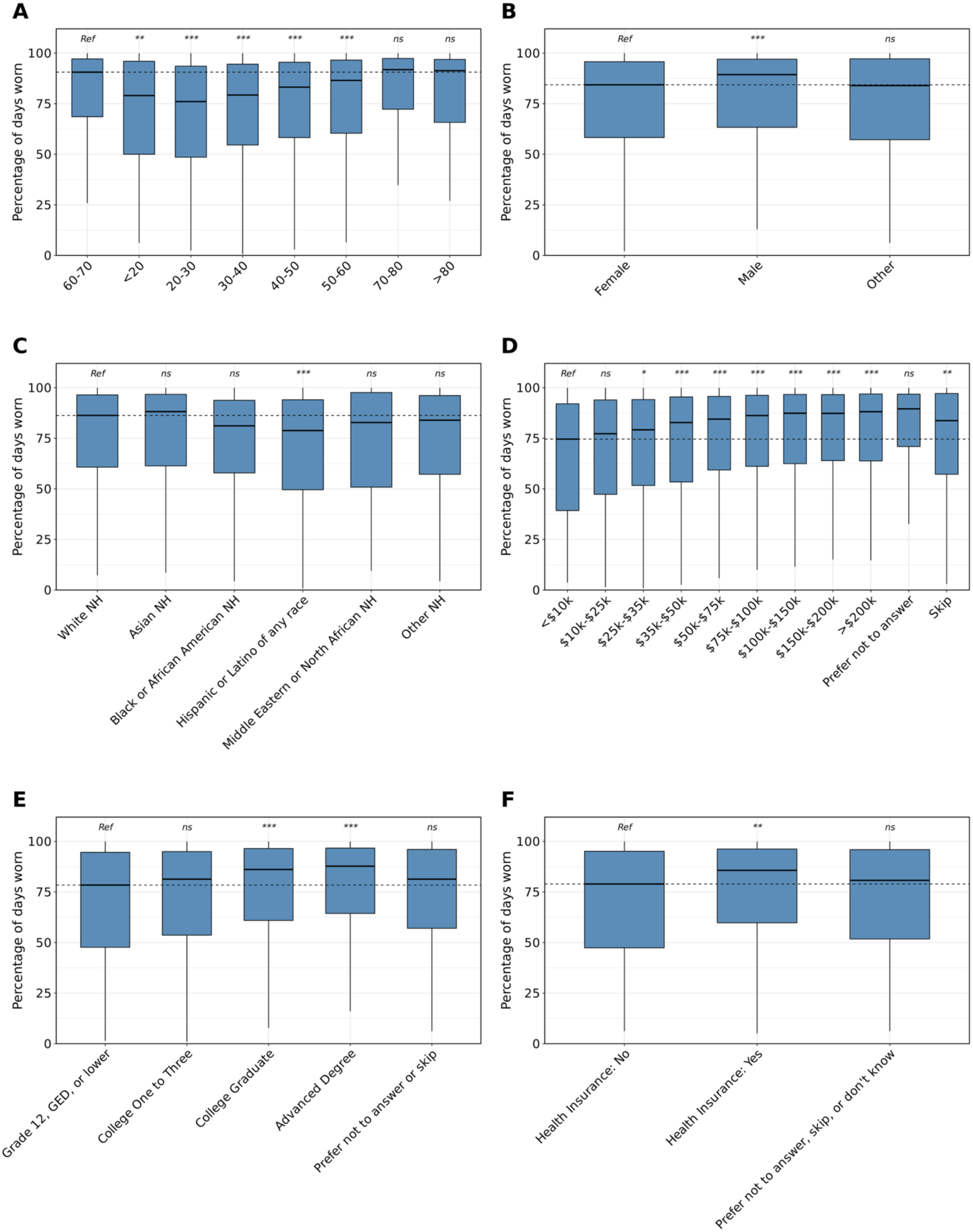
Demographic differences in the percentage of days individuals wear their Fitbit. NH=non-Hispanic; ME=Middle Eastern; NA=Northern African; *ns*=not significant, *=*P*<0.05, **=*P*<0.01, ***=*P*<0.001 after Bonferroni correction *The percentage of days individuals in the AoU cohort wore their Fitbit by age (A), sex (B), and race/ethnicity (C), annual income (D), highest level of education (E), and health insurance status (F). Data in each group were compared using linear regression and are expressed as mean and 95% confidence intervals. Statistical tests were run relative to a reference group represented as “Ref”. The dashed line indicates the median of the reference group. Individuals who were younger, female, Hispanic/Latino of any race, lower income, lower education level, and without health insurance wore their Fitbits a lower percentage of days relative to their control counterparts*.

We then aimed to compare variations in wearable device wear time among these groups, measured by the number of hours per day the device was worn. We found 1) individuals who were 20-30 wore their device fewer hours than those who were 60-70 (estimate=-0.68 hours, 95% CI=-0.89, – 0.47, *P*<0.001), 2) males wore their Fitbit more hours per day than females (estimate=0.68 hours, 95% CI=0.55, 0.81, *P*<0.001), and 3) individuals who were Black NH and Hispanic/Latino of any race wore their Fitbit fewer hours per day than those who were White NH (Black NH: estimate=-0.47 hours, 95% CI=-0.74, –0.20, *P*<0.01; Hispanic/Latino of any race: estimate=-0.49 hours, 95% CI=-0.73, –0.24, *P*<0.001; Figures 3A-3C). With regards to SDoH features, we showed 1) individuals with a higher income wore their device more hours per day, 2) individuals with an advanced degree wore their Fitbit more than those whose highest completed education was grade 12, GED, or lower, and 3) there was no difference in the number of hours per day individuals with and without health insurance wore their Fitbit (Figures 3D-3F). When evaluating hours per day of Fitbit wear time by smoking and alcohol use, we found no significant difference between individuals who smoked or drank compared to those who did not (Figures S2A and S2B).

**Figure 3:**
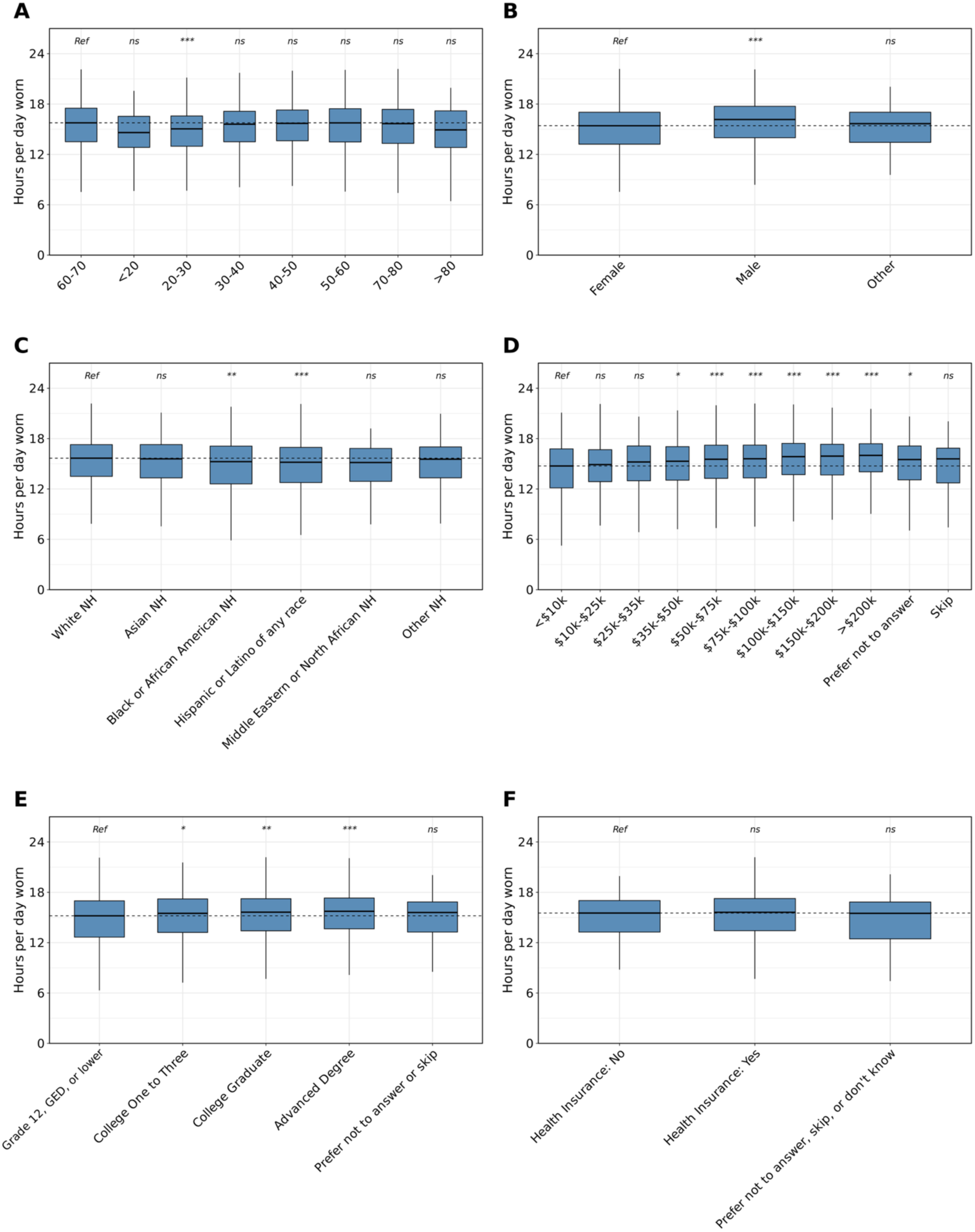
Demographic differences in the hours of daily Fitbit wear time. NH=non-Hispanic; ME=Middle Eastern; NA=Northern African; *ns*=not significant, *=*P*<0.05, **=*P*<0.01, ***=*P*<0.001 after Bonferroni correction *The number of hours per day individuals in the AoU cohort wore their Fitbit by age (A), sex (B), and race/ethnicity (C), annual income (D), highest level of education (E), and health insurance status (F). Data in each group were compared using linear regression and are expressed as mean and 95% confidence intervals. Statistical tests were run relative to a reference group represented as “Ref”. The dashed line indicates the median of the reference group. Individuals who were younger, female, Black NH, Hispanic and Latino of any race, lower income, and lower education wore their Fitbits fewer hours per day relative to their control counterparts*.

### Depressive, anhedonia, and anxiety symptoms are associated with reduced Fitbit wear time

Previous studies have suggested that mood and anxiety symptoms can influence factors related to user engagement, such as the amount of time individuals wear their wearable devices^21^. Therefore, it was hypothesized that those with depressive and/or anxiety symptoms would wear their device fewer days and fewer hours than those without. To probe this hypothesis, we used three questions from the survey data in AoU asking about one’s level of 1) depressive, 2) anxiety, and 3) anhedonia (the inability to feel pleasure or enjoyment from activities that are typically pleasurable) symptoms over the previous two week time period. We found that individuals with depressive symptoms wore their Fitbit a lower percentage of days and no difference in the hours per day than those without depressive symptoms (percentage: estimate=-6.00, 95% CI=-8.20, –3.81, *P*<0.001; hours: estimate=-0.25, 95% CI=-0.53, 0.02, *P*>0.05) (Figure 4A). The same pattern persisted in individuals with and without symptoms of anxiety (percentage: estimate=-3.74, 95% CI=-6.05, –1.43, *P*<0.01; hours: estimate=-0.08, 95% CI=-0.36, 0.20, *P*>0.05) (Figure 4A). Individuals with symptoms of anhedonia wore their Fitbit a fewer percentage of days and hours than those without anhedonia (percentage: estimate=-7.47, 95% CI=-9.67, –5.28, *P*<0.001; hours: estimate=-0.31, 95% CI=-0.58, –0.03, *P*<0.05)(Figure 4A). When analyzing multinomial survey responses with more granularity, we observed a dose-dependent trend with decreased percentage of wear time among those with more severe depressive, anxiety, and anhedonia symptoms; however there was no difference in the hours of wear time between those without any symptoms and those with any symptoms (Figure 4B). Lastly, given the common co-occurrence of depressive and anxiety symptoms, we directly compared the percentage and hours of wear time among individuals with depressive symptoms alone, anxiety symptoms alone, or both symptoms together compared to those with neither symptom group. The results demonstrated that individuals with depressive symptoms wore their Fitbit fewer days than those with neither symptom group (estimate=-7.22%, 95% CI=-12.6, –1.87, *P*<0.05), those with both symptoms wore their device significantly fewer days than those with neither symptom group (estimate=-6.70%, 95% CI=-9.35, –4.04, *P*<0.001), and there was no difference in the hours of wear time between those with depressive, anxiety, or both symptoms compared to those without any symptoms (Figure 4C).

**Figure 4:**
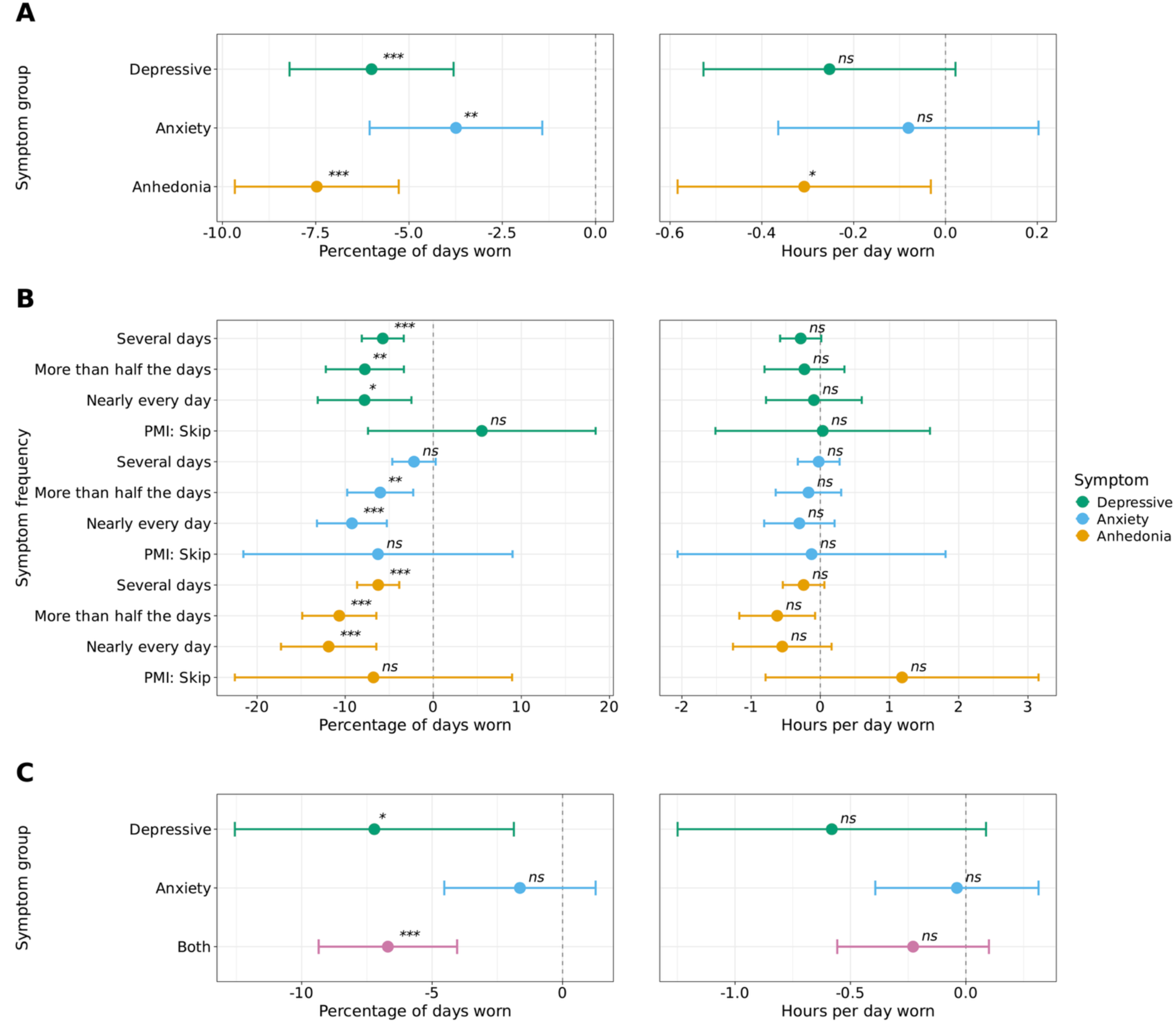
Differences in Fitbit wear time in individuals with depressive, anxiety, and anhedonia symptoms. *ns*=not significant, *=*P*<0.05, **=*P*<0.01, ***=*P*<0.001 after Bonferroni correction *The percentage of days (left) and hours per day (right) individuals in the AoU cohort wore their Fitbit by separate binary comparisons of individuals with versus without each symptom type (depressive [2,672 with and 3,542 without], anxiety [3,894 with and 2,320 without], or anhedonia [2,660 with and 3,554 without]) (A), symptom frequency (multinomial responses) (B), and depressive and anxiety symptoms in combination and isolation (C). The percentage of days and hours per day Fitbit devices were worn were analyzed using linear regression. All models adjusted for age, sex, race/ethnicity, annual income, education level, and health insurance status. The dashed line indicates the reference group (no symptoms for that comparison). Data are expressed as mean and 95% confidence intervals. Individuals with depressive or anhedonia symptoms consistently wore their devices fewer days, while the relationship with anxiety symptoms appeared more complex and potentially influenced by symptom comorbidity*.

### Individuals with depressive and anxiety diagnoses wear their fitbit less than those with neither depressive nor anxiety diagnoses

To better elucidate the association between depression and anxiety with Fitbit wear time, we leveraged the AoU EHR data to generate a cohort of individuals diagnosed with major depressive disorder (MDD) and/or anxiety disorder (AD). First, we compared the percentage and hours of daily Fitbit wear time amongst those with and without a diagnosis of MDD and found that those with MDD wore their Fitbit less than those without MDD (percentage: estimate=-59.7, 95% CI=-61.0, –58.4, *P*<0.001; hours: estimate=-9.59, 95% CI=-9.85, –9.33, *P*<0.001; Figure 5A). Similar findings were observed among those with AD (percentage: estimate=-56.5, 95% CI=-57.7, –55.2, *P*<0.001; hours: estimate=-9.03, 95% CI=-9.27, –8.78, *P*<0.001; Figure 5A). Consistent with our methods with depressive and anxiety symptoms in the survey data, we combined and assessed the relationship between Fitbit wear time among individuals with 1) both MDD and AD, 2) MDD only, 3) AD only, and 4) neither. The findings showed that individuals in groups 1, 2, and 3 wore their Fitbit significantly fewer days and hours per day than those with neither MDD nor AD (group 4; Figure 5B).

**Figure 5:**
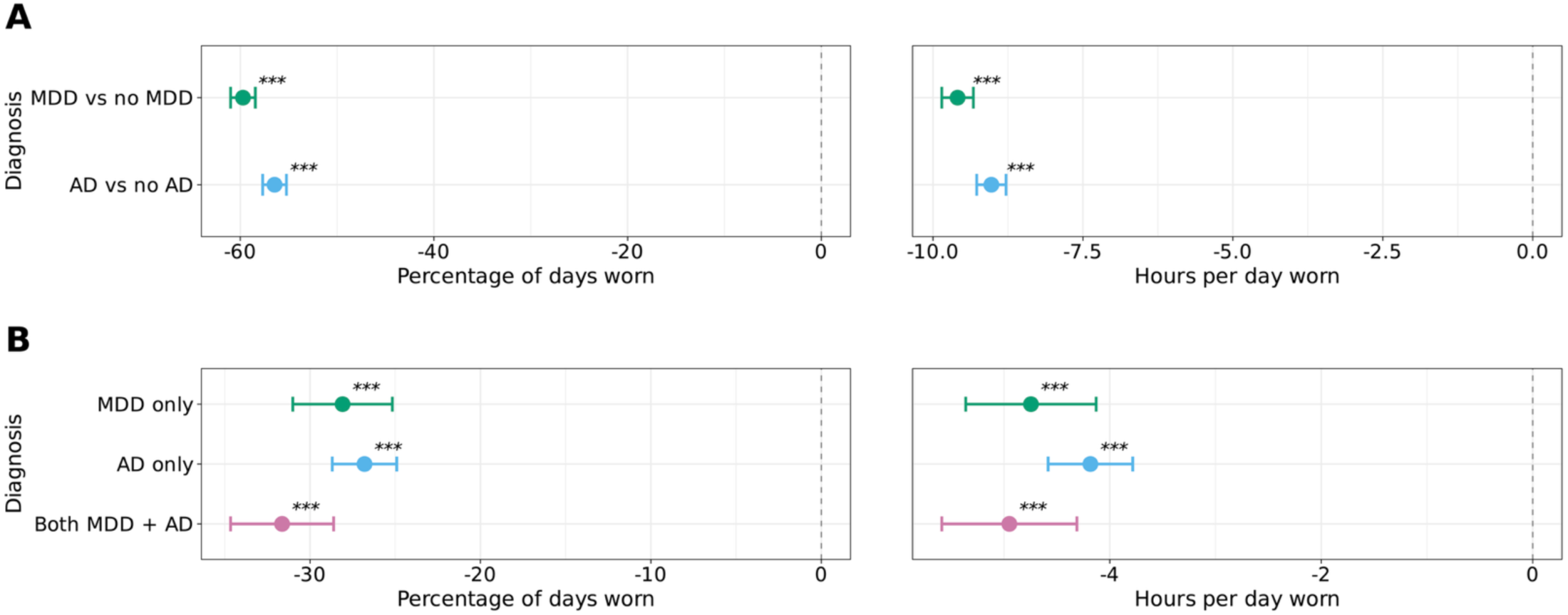
Differences in Fitbit wear time in individuals with depression and anxiety diagnoses. MDD=major depressive disorder; AD=anxiety disorder; *ns*=not significant, *=*P*<0.05, **=*P*<0.01, ***=*P*<0.001 after Bonferroni correction *The percentage of days (left) and hours per day (right) individuals in the AoU cohort wore their Fitbit by a diagnosis of depression (1,364 with and 10,425 without) or anxiety (1,830 with and 9,926 without without) (A) or combined depression and anxiety (215 with depression only, 528 with anxiety only, 201 combined, and 9,533 with neither) (B). The percentage of days and hours per day Fitbit devices were worn were analyzed using linear regression. All models adjusted for age, sex, race/ethnicity, annual income, education level, and health insurance status. Data are expressed as mean and 95% confidence intervals. The dashed line indicates the reference group (no symptoms for that comparison). Individuals with diagnosed MDD and AD wore their Fitbits fewer days and hours per day than controls, with comorbid MDD and AD showing further reductions in both measures*.

### Mental disorder diagnoses show disorder-specific associations with Fitbit wear time

To expand upon our results of wearable wear time amongst those with MDD or AD, we wondered whether decreased Fitbit wear time would persist among individuals with other types of mental disorder diagnoses. Mental disorders were categorized using phecodes (Table S1), with 3,335 (28.0%) individuals having at least one mental disorder. We first examined whether there was a difference in the percentage of days individuals with each mental disorder wore their Fitbit compared to those without any mental disorder. Our results showed individuals with mood disorders, anxiety disorders, and tobacco use disorder wore their Fitbit a fewer percentage of days and hours than those without any mental disorder, while those with neurological disorders wore their Fitbit a higher percentage of days and hours compared to those without any mental disorder (Figure 6).

**Figure 6:**
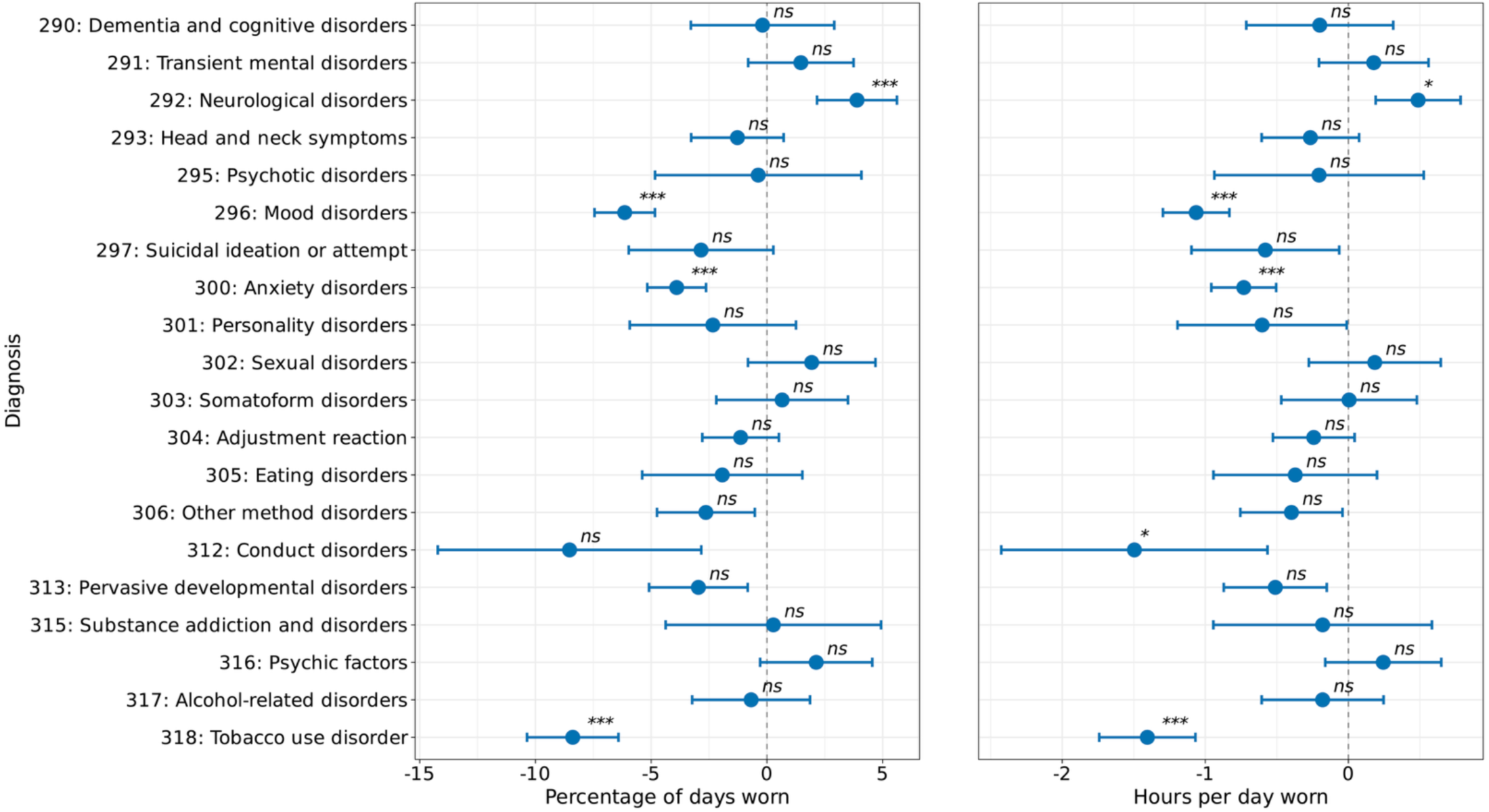
Association between mental disorder categories and Fitbit wear time. *ns*=not significant, *=*P*<0.05, **=*P*<0.01, ***=*P*<0.001 after Bonferroni correction *The percentage of days (left) and hours per day (right) individuals in the AoU cohort wore their Fitbit by various phecode-categorized mental disorders. The percentage of days and hours per day Fitbit devices were worn was analyzed using linear mixed-effects models, with person ID included as a random effect. All models adjusted for age, sex, race/ethnicity, annual income, education level, and health insurance status. Data are expressed as mean and 95% confidence intervals. The dashed line indicates the reference group (no mental disorders). Individuals with mood disorders and other mental disorders wore their Fitbit the fewest percentage of days and hours per day than their control counterparts*.

To examine Fitbit wear time more closely with the incidence of each mental disorder, we aimed to determine whether the percentage of Fitbit wear time differed within individuals after they were diagnosed with a mental disorder compared to before their diagnosis. To answer this question, we calculated and compared the percentage of days each individual wore their Fitbit before and after the date of diagnosis. The same analysis was performed in the AoU cohort without any diagnosed mental disorders as a control. Among individuals without a mental disorder (controls), there was a decrease in wear time after the index date (mean estimate=-7.74, 95% CI=-14.68, –0.80, *P*<0.05) (Figure S3). Compared to controls, there was no significant difference in the percentage of days or hours per day worn after the index date among individuals with any type of mental disorder (Figure S3). While we did observe individuals with conduct disorders exhibited a larger decline in wear time percentage after diagnosis compared to controls, this was not significant after Bonferroni correction (interaction: –19.9%, *P*=0.03, *ns* after Bonferroni correction) (Figure S3). Examining hours per day worn, individuals without mental disorders showed no significant change after the index date (estimate=-0.76 hours, *P*>0.05) (Figure S3). Additionally, no diagnostic groups differed significantly from controls in baseline wear time (all main effects *P*>0.05) (Figure S3).

### Chronic conditions show disease-specific associations with Fitbit wear time

We examined whether there existed different patterns of Fitbit wear time by classes of disease compared to healthy controls (i.e., individuals without any Phecode-grouped recorded disorders). Disease classes were categorized using phecodes (Table S2), with 6,161 (52.0%) individuals having at least one chronic disease. We found that those with blood disorders, dermatologic disorders, infectious diseases, injuries and poisoning, mental disorders, musculoskeletal disorders, neoplasms, conditions during the perinatal period, and symptoms groups wore their Fitbit significantly more than healthy controls (note the control group in this analysis is healthy controls as opposed to individuals without any mental disorder in the prior analysis) (Figure 7). When comparing hours of wear time, we found that individuals with blood disorders, infectious diseases, and conditions during the perinatal period wore their Fitbit significantly more than healthy controls (Figure 7).

**Figure 7:**
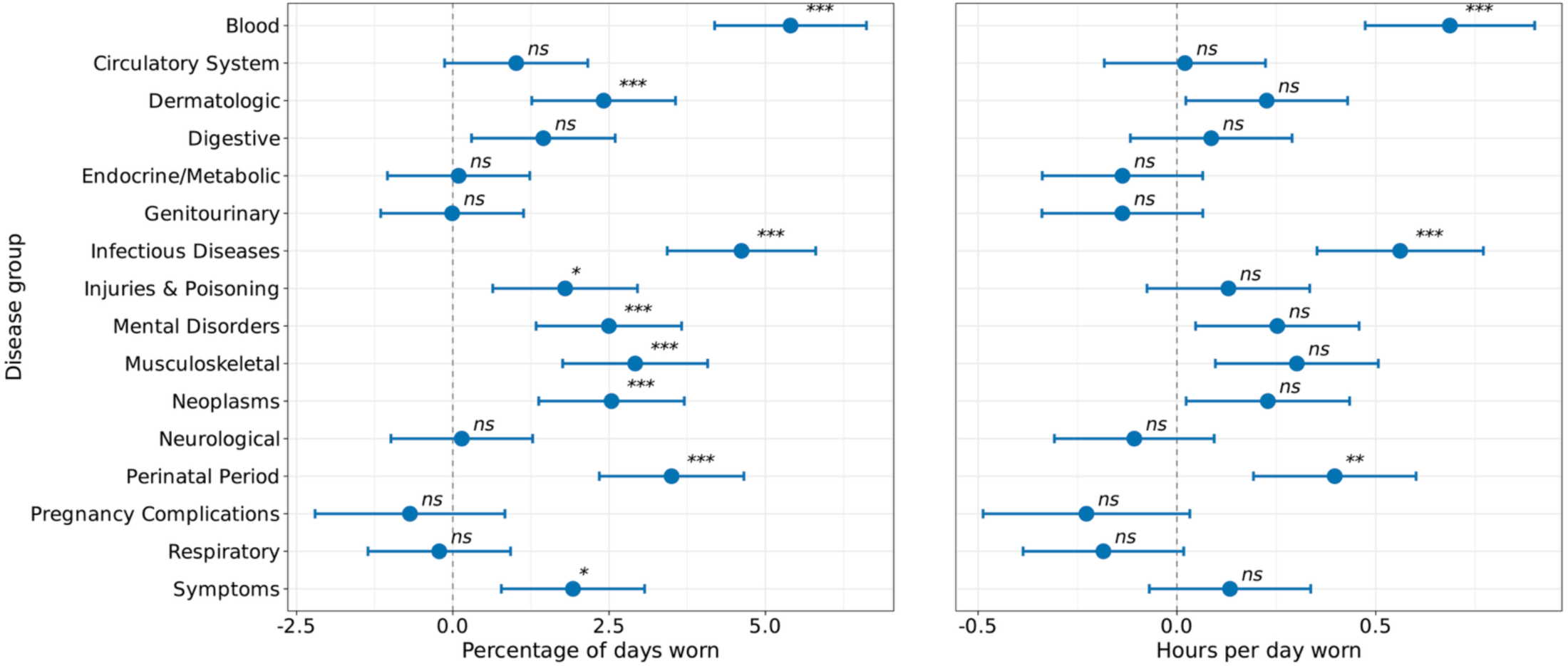
Association between chronic disease categories and Fitbit wear time. *ns*=not significant, *=*P*<0.05, **=*P*<0.01, ***=*P*<0.001 after Bonferroni correction *The percentage of days (left) and hours per day (right) individuals in the AoU cohort wore their Fitbit by disease groups. The percentage of days and hours per day Fitbit devices were worn was analyzed using linear mixed-effects models, with person ID included as a random effect. All models adjusted for age, sex, race/ethnicity, annual income, education level, and health insurance status. Data are expressed as mean and 95% confidence intervals. The dashed line indicates the reference group (healthy controls). Individuals with blood disorders and infectious diseases wore their Fitbit the highest percentage of days and hours per day than their control counterparts*.

Similar to our prior analysis, we compared the percentage of wear time before and after the diagnosis of each class of disorders. The control group (individuals without chronic conditions) demonstrated a 5.16% decline in wear time after the index date (95% CI=-6.31, –4.01, *P*<0.001), consistent with natural temporal trends in device engagement (Figure S4). After the index date, individuals with genitourinary disorders (estimate=-3.19%, 95% CI=-4.74, –1.65, *P*<0.01, estimate=-0.51 hours, 95% CI=-0.77, –0.25, *P*<0.01), injury and poisoning (estimate=-3.34%, 95% CI=-5.03, – 1.66, *P*<0.01, estimate=-0.64, 95% CI=-0.93, –0.36, *P*<0.001), mental disorders (estimate=-0.42 hours, 95% CI=-0.69, –0.14, *P*<0.05), musculoskeletal disorders (estimate=-2.61%, 95% CI=-4.15, –1.06, *P*<0.05, estimate=-0.47 hours, 95% CI=-0.73, –0.21, *P*<0.01), and respiratory disorders (estimate=-3.43%, 95% CI=-4.97, –1.90, *P*<0.001; estimate=-0.59 hours, 95% CI=-0.85, –0.33, *P*<0.001) all wore their device fewer hours per day than controls (Figure S4).

### Wear time handling methods reveal differential bias and data retention across analytical approaches

As a practical application, we demonstrated how different analytical approaches to handling wear time affect the estimation of a well-studied clinical relationship: daily step count differences between individuals with and without major depressive disorder (MDD), which was selected because reduced physical activity is a well-documented feature of depression. Current practice in wearable device research commonly addresses wear time variability by excluding ‘subthreshold’ days that fail to meet minimum wear criteria; however, if non-compliance is non-random and associated with the condition under study, this exclusion introduces selection bias that could distort clinical effect estimates. We compared five methodological approaches: 1) the standard compliance threshold (≥10 hours of wear time, >100 steps, and <45,000 steps); 2) covariate adjustment, using all days of data while including hours of daily wear time as a covariate; 3) metric normalization, calculating steps per hour of wear time rather than total daily steps; 4) propensity score matching based on hours of wear time at the person-day level; and 5) adaptive thresholds, systematically evaluating minimum wear time thresholds from 1 to 10 hours.

Across all five approaches, individuals with MDD took fewer steps than those without MDD, though the magnitude and statistical significance of the difference varied by method (Figures 8A and 8B). The standard compliance threshold yielded the largest estimated group difference (999 steps/day, 95% CI=727, 1,271, *P*<0.001), while covariate adjustment produced a smaller but highly significant difference (796 steps/day, 95% CI=551, 1,041 *P*<0.001), suggesting that a portion of the apparent activity gap under standard filtering reflects unaccounted wear time variation rather than true differences in physical activity (Figure 8A). Metric normalization confirmed reduced activity among those with MDD on a rate-adjusted scale (55 fewer steps/hour, 95% CI=34, 76, *P*<0.001; Figure 8A). Interestingly, propensity score matching produced an estimate in the same direction but was not statistically significant (402 fewer steps/day, 95% CI=-25, 828, P=0.065), which may be attributed to severe data loss from day-level matching (93% of data excluded), which substantially reduced statistical power and selected an unrepresentative subset of low-wear-time control days (Figure 8A). Adaptive threshold analysis revealed that the MDD versus no MDD step difference remained significant across all thresholds from 1 to 10 hours (all *P*<0.001), and critically, that wear time remained significantly different between groups at every threshold, including 10-hours (*P*<0.001; Figure 8B). This finding demonstrates that compliance filtering alone does not control for residual confounding wear time differences between clinical populations.

**Figure 8:**
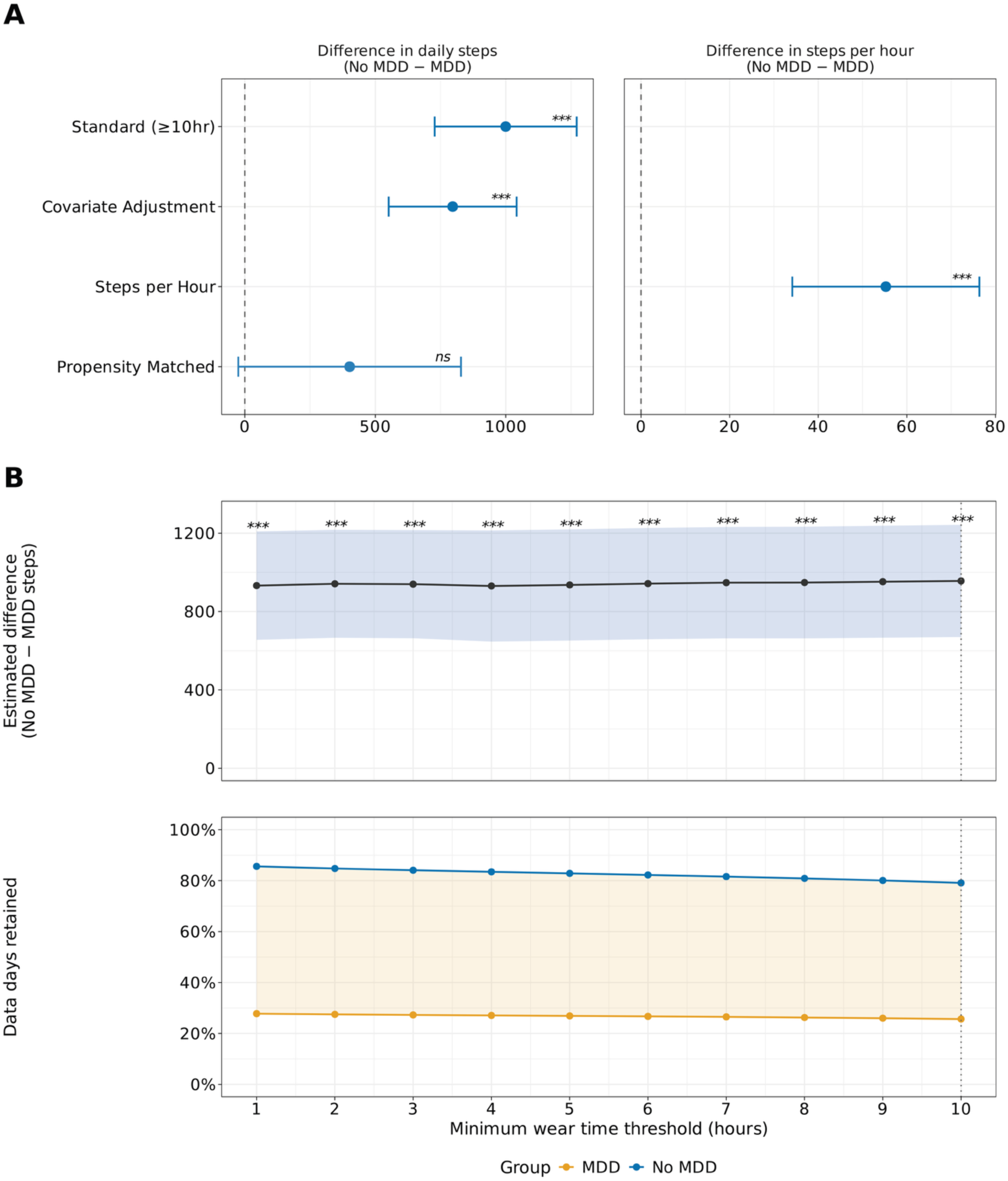
Comparison of analytical methods for estimating daily step count differences between individuals with and without MDD. MDD=major depressive disorder, *ns*=not significant, *=*P*<0.05, **=*P*<0.01, ***=*P*<0.001 *Estimated marginal mean daily steps and group differences across five analytical approaches for handling Fitbit wear time in individuals with and without major depressive disorder (MDD). (A) Estimated group differences (No MDD − MDD) in daily steps across analytical approaches: standard compliance filtering (≥10 hours of wear time), covariate adjustment for wear time, and propensity score matching on wear time, with the steps per hour metric normalization approach shown separately on the right. (B) Estimated group difference (No MDD − MDD) in daily steps across adaptive minimum wear time thresholds from 1 to 10 hours, with the shaded region indicating 95% confidence intervals (top); Percentage of data days retained by group across minimum wear time thresholds, with the dotted line indicating the standard 10-hour threshold (bottom). All models were linear mixed-effects models with person ID as a random effect, adjusted for age, sex, and race/ethnicity. Data are expressed as estimated marginal means and 95% confidence intervals. The group difference in daily steps remained significant across all analytical methods and wear time thresholds except propensity score matching, which showed a similar direction of effect but did not reach statistical significance (P=0.065). Individuals with MDD consistently took fewer daily steps than those without MDD regardless of the wear time handling approach used*.

Each method retained different amounts of data, with important implications for statistical power and representativeness. The standard compliance threshold retained 485,232 days (72.8% of total data), but this filtering disproportionately excluded data from the MDD cohort: 58,732 days (74.4%) of MDD data were removed compared to 122,936 days (20.9%) from the no MDD cohort (Table 3). Metric normalization retained 525,212 days (78.8%), excluding only days with zero recorded wear time (Table 3). Propensity score matching retained just 47,520 days (7.1%), with the severe data loss driven by 1:1 day-level matching against a control pool roughly 22 times larger than the MDD group, producing an unrepresentative subset that substantially reduced statistical power (Table 3). Covariate adjustment retained all 666,900 days (100%), including an additional 181,668 days that would have been discarded under the standard threshold (Table 3). Adaptive threshold analysis further demonstrated that filtering disproportionately excluded MDD data at every threshold from 1 to 10 hours (Figure 8B).

**Table 3:**
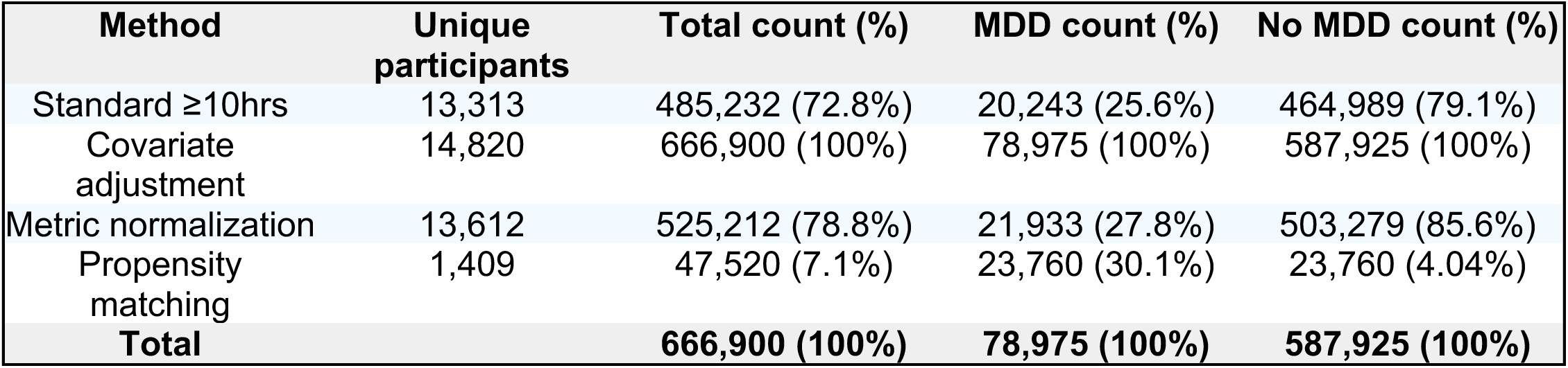
Standard wear time criteria exclude more days of Fitbit data from individuals with MDD.

| **Method** | **Unique participants** | **Total count (%)** | **MDD count (%)** | **No MDD count (%)** |
| --- | --- | --- | --- | --- |
| Standard $\geq 10$hrs | 13,313 | 485,232 (72.8%) | 20,243 (25.6%) | 464,989 (79.1%) |
| Covariate adjustment | 14,820 | 666,900 (100%) | 78,975 (100%) | 587,925 (100%) |
| Metric normalization | 13,612 | 525,212 (78.8%) | 21,933 (27.8%) | 503,279 (85.6%) |
| Propensity matching | 1,409 | 47,520 (7.1%) | 23,760 (30.1%) | 23,760 (4.04%) |
| **Total** |  | **666,900 (100%)** | **78,975 (100%)** | **587,925 (100%)** |

## Discussion

Wearable devices present transformative opportunities for personalized healthcare through continuous monitoring of digital biomarkers; however, our findings demonstrate that individual variations in device wear time represent a critical and previously undercharacterized source of methodological bias in biomedical wearable device research. Using Fitbit data from approximately 11,000 participants in the *All of Us* Research Program, we conducted the first large-scale systematic assessment of wear time across demographics, social determinants of health, lifestyle factors, mental health symptoms, and disease, revealing that wear time was non-randomly distributed across populations: it was higher among males and increased with age, income, and education, but decreased with depressive, anxiety, and anhedonia symptoms, with reductions more pronounced following clinical diagnoses compared to symptom-based classifications. Individuals with chronic conditions displayed differential levels of wear time compared to healthy controls. Critically, we demonstrate that the widely used ≥10-hour daily compliance threshold can disproportionately exclude data from disease populations, with 74.4% of data days excluded among individuals with major depressive disorder compared to 20.9% for controls. To address this, we propose and empirically evaluated a flexible methodological framework, including covariate adjustment, metric normalization, propensity score matching, and adaptive thresholds, that can be applied individually or in combination to optimize data retention while maintaining analytical rigor. These findings establish wear time as both a methodological consideration that must be addressed in wearable device research design and a potential informative signal in its own right.

Several factors are known to impact wearable device wear time, including age, education, income, occupation, fitness levels, and health conditions^38^. We observed that males, older adults, and those with higher income and education status tended to wear their Fitbit more than their counterparts. Although younger adults demonstrate higher digital health literacy and wearable adoption rates^39^, among individuals who have already adopted a device, older adults wore their Fitbit more consistently, consistent with previous studies^40^. Previous research suggests that wearable devices support “active aging”, defined as enhancing quality of life throughout the aging process^41^. Qualitative analysis indicated older adults are motivated to learn about their health, highlighting a key value of wearables. Research also suggests that older adults with specific needs, such as fall risk, are more likely to engage with their devices as a result of that motivation^42,43^. We were also surprised to find males displayed higher wear time than females despite the AoU cohort being predominantly female (70%). This may reflect self-selection bias, where males who chose to enroll in a research study and share their Fitbit data represent a more health-motivated and physically active subgroup than the general male population, a pattern consistent with research showing males who engage with wearable device features tend to be more physically active than their female counterparts^44^. While the difference was statistically significant, the absolute magnitude was very small. Notably, this finding contrasts with self-reported data suggesting women are more likely to use wearable health devices than men^45^, further supporting the idea that the bring-your-own-device enrollment model may have introduced motivational self-selection that disproportionately filtered the male sample^46,47^.We also observed individuals with higher incomes and education levels engage more with wearable devices, consistent with research demonstrating that children from families with higher education and income show greater device adherence^48^. This may be due to systemic factors, where individuals of lower socioeconomic status prioritize more immediate needs over wearable technology, limiting their device wear time and preventing them from fully experiencing its benefits. Biases in wearable device deployment and data collection exist^3,49–51^ and structural and systemic barriers, such as concerns about data privacy, accuracy, language barriers, and access to health care technology, impact use of wearables among racial and ethnic minorities^46,47,52–55^. Recognizing these disparities underscores the importance of both equitable device deployment and analytical approaches that account for wear time variability, ensuring data from lower socioeconomic populations is preserved rather than systematically excluded through strict compliance criteria^47^.

Our findings also showed individuals with depressive, anxiety, and anhedonia symptoms exhibited lower levels of wearable device wear time compared to those without symptoms. This is consistent with prior work suggesting depressive and anxiety symptoms are linked to lower healthy lifestyle scores, which may contribute to decreased motivation for health-tracking behaviors^21,56^. Lower levels of wear time were more pronounced in individuals with diagnosed depression and anxiety disorders compared to those reporting symptoms without formal diagnoses, likely reflecting greater symptom severity in the diagnosed group. Wear time was consistently reduced in those with any diagnosed mental disorder compared to those without, with notable variation across specific disorders: individuals with tobacco use disorder showed lower wear time, consistent with broader patterns of reduced health-promoting behaviors and sedentary lifestyle in this population^57^, while those with neurological disorders showed higher wear time, potentially reflecting the particular clinical utility of continuous device monitoring in this group^58^. Expanding this analysis to chronic disorders more broadly revealed similarly variable patterns; while detailed interpretation of individual conditions is beyond the scope of this paper, future research should explore specific disorders in greater depth, considering factors such as disease severity and condition-specific characteristics to better contextualize wear time patterns.

When examining differences in wear time across individuals with and without MDD in our example, we observed that those with MDD were disproportionately impacted by filtering on compliance, with nearly 75% of days of data being excluded, compared to those without MDD who only had about 21% of days excluded (which is still sizeable). This disproportionate data loss has critical implications for statistical analyses and machine learning models. By incorporating hours of wear time as a covariate rather than applying strict compliance filters, we can rescue these otherwise excluded days of data. This approach not only increases statistical power, particularly important for chronic conditions disproportionately affected by wear time, but also reduces selection bias by retaining data from vulnerable populations who would otherwise be systematically excluded from analyses. Importantly, the two methods produce different absolute activity estimates where compliant-only data yields higher step counts because low-activity days are disproportionately excluded, a consideration for downstream clinical and public health applications where accurate activity estimates are needed. Our results demonstrate that this methodological shift enables more inclusive and representative digital health research while maintaining analytical rigor.

We acknowledge that the ≥10-hour wear time threshold has sound methodological origins and remains useful for specific applications, such as estimating representative daily activity in healthy populations^18^. However, for disease-specific research, we propose that reduced wear time is often informative rather than simply indicative of invalid data. Several methodological approaches can address variability in wear time while preserving these observations. When the primary goal is maximizing statistical power and sample representativeness, including hours of wear time as a covariate retains all available data while statistically controlling for differential wear across groups, making it particularly suited for group-level comparisons of activity outcomes where data loss would disproportionately affect one group^59^. An additional consideration is the timing of available data, as systematic missingness during specific hours can attenuate or exaggerate circadian patterns. When the outcome of interest is an activity metric that scales directly with wear duration, such as step counts, normalizing by wear time (e.g., steps per hour) provides an intuitive rate-based measure that is inherently comparable across participants with variable wear, though this approach assumes a relatively constant activity rate throughout the day and requires a minimum wear time floor to avoid unstable estimates^59^. When the analytical design requires equivalent device exposure between comparison groups, such as in case-control studies, propensity score matching on wear time ensures balanced distributions prior to analysis, though this comes at the cost of reduced sample size and generalizability, as demonstrated by the substantial data loss observed in our MDD example^60^. When researchers seek to retain more data than the standard threshold while still ensuring a minimum level of data quality per day, an adaptive threshold calibrated to the study population can balance data retention with analytical validity, particularly when wear time differences between groups diminish at lower thresholds^61^. These approaches are not mutually exclusive; for example, a researcher could apply an adaptive threshold to ensure minimum data quality and then include residual wear time variation as a covariate. The optimal combination will depend on the research question, outcome of interest, and study population, and we encourage researchers to report results under multiple approaches as a form of sensitivity analysis. Future studies comparing these approaches against gold-standard activity measures (e.g., direct observation) would help establish validity benchmarks for different research contexts. By offering a flexible methodological framework rather than a single prescriptive solution, we aim to equip researchers with practical tools for maximizing wearable device data retention while maintaining analytical rigor across diverse clinical populations (Table 1).

Although this study is one of the first to characterize large-scale, biomedical wearable device wear time, it is not without limitations. For example, EHR-based analyses may not accurately capture disease incidence depending on where and when data are entered, as symptoms often precede both diagnosis and the subsequent recording in the EHR^62,63^. For this reason, we excluded certain condition groups where the first recorded date was known to be inaccurate or inappropriate (e.g., congenital disorders). Nonetheless, EHR data are still commonly used to estimate incidence, and our results should be interpreted with this limitation in mind^62,63^. Second, the analysis performed in this AoU cohort (v7) used a bring-your-own-device model, meaning individuals who already owned their Fitbit consented to share their data. This results in a demographic imbalance of individuals who own wearable devices, which tends to favor those of higher socioeconomic status^47^. Since the time of analysis, AoU updated their data to v8, which contains individuals from the WEAR study, which gave individuals from underrepresented communities a Fitbit and resulted in a new total sample size of approximately 60,000 individuals with Fitbit data in AoU^64^. It would be valuable for future work to replicate this study in a more heterogeneous cohort. Further, there was variability in the sample size in specific disease groups, which may limit the generalizability of our results. However, this is a common limitation of retrospective observational studies that are not collecting data for any particular population and allows us to conduct analyses across several diseases. Third, AoU has since added new Fitbit metadata, including device type and approximate battery level, which may influence wear time patterns and warrant investigation in future analyses.^65^. Finally, the calculation used to estimate hours of wear time, which was based on published literature^4,25^, relies on counting the number of non-zero steps, which may underestimate wear time by excluding periods when the individual was sedentary, although it is unusual for step counts to be exactly zero in sedentary individuals due to small movements. Future analyses could consider distinguishing between missing data (NAs) and minutes with recorded zero steps, or alternatively, use the presence of intraday heart rate measurements as an indicator of wear time^40^.

Overall, wear time with wearable devices varies across individuals based on a range of demographic and personal health factors. Wear time-related information can not only offer insights and serve as additional features alongside signals from wearable sensors (e.g., heart rate, physical activity, and sleep), but it can also be leveraged to maximize the amount of wearable device data available for analysis in statistical and machine learning models. Moreover, the exclusion of data based on insufficient wear time creates systematic biases, as data loss disproportionately affects specific sociodemographic and disease groups. Implementing appropriate methods and incorporating wear time data in analyses can enhance the utility and richness of longitudinal wearable device data, reinforcing the role of these devices in advancing equitable and personal health.

## Acknowledgments

The *All of Us* Research Program would not be possible without the partnership of its participants. The *All of Us* Research Program is supported by the National Institutes of Health, Office of the Director: Regional Medical Centers: 1 OT2 OD026549; 1 OT2 OD026554; 1 OT2 OD026557; 1 OT2 OD026556; 1 OT2 OD026550; 1 OT2 OD 026552; 1 OT2 OD026553; 1 OT2 OD026548; 1 OT2 OD026551; 1 OT2 OD026555; IAA #: AOD 16037; Federally Qualified Health Centers: HHSN 263201600085U; Data and Research Center: 5 U2C OD023196; Biobank: 1 U24 OD023121; The Participant Center: U24 OD023176; Participant Technology Systems Center: 1 U24 OD023163; Communications and Engagement: 3 OT2 OD023205; 3 OT2 OD023206; and Community Partners: 1 OT2 OD025277; 3 OT2 OD025315; 1 OT2 OD025337; 1 OT2 OD025276.

## Data availability

Access to the All of Us Research Program (AoURP) dataset is governed by the program’s data security and privacy policies. Eligible investigators who have completed AoURP’s required training and registered on the Researcher Workbench may request study data and code from the corresponding author.

## Author contributions

EH and MAH conceived the study. EH and MAH designed the study. EH and EC analyzed the data. EH, EC, MS, HM, HH, ZB, JD, and MAH interpreted the results. EH wrote the draft manuscript. MAH provided supervision and acquired funding. All authors reviewed and edited the final manuscript for publication.

## References

1. Dunn, J., Runge, R. & Snyder, M. Wearables and the medical revolution. Per. Med. 15, 429–448 (2018).

2. Yetisen, A. K., Martinez-Hurtado, J. L., Ünal, B., Khademhosseini, A. & Butt, H. Wearables in medicine. Adv. Mater. 30, e1706910 (2018).

3. Seneviratne, S. et al. A survey of wearable devices and challenges. IEEE Commun. Surv. Tutor. 19, 2573–2620 (2017).

4. Master, H. et al. Association of step counts over time with the risk of chronic disease in the All of Us Research Program. Nat. Med. 28, 2301–2308 (2022).

5. Zheng, N. S. et al. Sleep patterns and risk of chronic disease as measured by long-term monitoring with commercial wearable devices in the All of Us Research Program. Nat. Med. 30, 2648–2656 (2024).

6. Mishra, T. et al. Pre-symptomatic detection of COVID-19 from smartwatch data. *Nat*. Biomed. Eng. 4, 1208–1220 (2020).

7. Radin, J. M. et al. Long-term changes in wearable sensor data in people with and without Long Covid. NPJ Digit. Med. 7, 246 (2024).

8. Grzesiak, E. et al. Assessment of the feasibility of using noninvasive wearable biometric monitoring sensors to detect influenza and the common cold before symptom onset. *JAMA Netw*. Open 4, e2128534 (2021).

9. Ballinger, B., et al. DeepHeart: Semi-Supervised Sequence Learning for Cardiovascular Risk Prediction. AAAI https://aaai.org/papers/11891-deepheart-semi-supervised-sequence-learning-for-cardiovascular-risk-prediction/ (2023).

10. Perez, M. V. et al. Large-scale assessment of a smartwatch to identify atrial fibrillation. N. Engl. J. Med. 381, 1909–1917 (2019).

11. Bent, B. et al. Non-invasive wearables for remote monitoring of HbA1c and glucose variability: proof of concept. BMJ Open Diabetes Res. Care 9, e002027 (2021).

12. Lehmann, V. et al. Noninvasive hypoglycemia detection in people with diabetes using smartwatch data. Diabetes Care 46, 993–997 (2023).

13. Jeong, H., Kim, H., Kim, R., Lee, U. & Jeong, Y. Smartwatch wearing behavior analysis: A longitudinal study. Proc. ACM Interact. Mob. Wearable Ubiquitous Technol. 1, 1–31 (2017).

14. Ogbanufe, O. & Gerhart, N. Watch it! Factors driving continued feature use of the smartwatch. Int. J. Hum. Comput. Interact. 1–16 (2017).

15. Visuri, A. et al. Understanding usage style transformation during long-term smartwatch use. Pers. Ubiquitous Comput. 25, 535–549 (2021).

16. Saheb, T., Cabanillas, F. J. L. & Higueras, E. The risks and benefits of Internet of Things (IoT) and their influence on smartwatch use. Span. J. Mark.-ESIC 26, 309–324 (2022).

17. Siepmann, C. & Kowalczuk, P. Understanding continued smartwatch usage: the role of emotional as well as health and fitness factors. Electron. Mark. 31, 795–809 (2021).

18. Troiano, R. P. et al. Physical activity in the United States measured by accelerometer. Med. Sci. Sports Exerc. 40, 181–188 (2008).

19. Beauchamp, U. L., Pappot, H. & Holländer-Mieritz, C. The use of wearables in clinical trials during cancer treatment: Systematic review. JMIR MHealth UHealth 8, e22006 (2020).

20. Nyrop, K. A. et al. Measuring and understanding adherence in a home-based exercise intervention during chemotherapy for early breast cancer. Breast Cancer Res. Treat. 168, 43–55 (2018).

21. Pathiravasan, C. H. et al. Factors associated with long-term use of digital devices in the electronic Framingham Heart Study. NPJ Digit. Med. 5, 195 (2022).

22. All of Us Research Program Investigators et al. The ‘All of Us’ Research Program. N. Engl. J. Med. 381, 668–676 (2019).

23. Hurwitz, E., et al. Unlocking the potential of wear time of a wearable device to enhance postpartum depression screening and detection: Cross-sectional study. JMIR Form. Res. 9, e67585 (2025).

24. Bates, D., Mächler, M., Bolker, B. & Walker, S. Fitting Linear Mixed-Effects Models using lme4. arXiv [stat.CO] (2014).

25. Hurwitz, E. et al. Harnessing consumer wearable digital biomarkers for individualized recognition of postpartum depression using the All of us research program data set: Cross-sectional study. JMIR MHealth UHealth 12, e54622 (2024).

26. Bland, J. M. & Altman, D. G. Multiple significance tests: the Bonferroni method. BMJ 310, 170 (1995).

27. Data and Statistics Dissemination Policy. Preprint at https://www.researchallofus.org/wp-content/themes/research-hub-wordpress-theme/media/2020/05/AoU_Policy_Data_and_Statistics_Dissemination_508.pdf.

28. COVID-19 Participant Experience (COPE) Survey. Preprint at https://databrowser.researchallofus.org/assets/surveys/COPE_survey_June_2020_English.pdf.

29. PheKB. https://phekb.org/phenotype/depression.

30. Zheng, N. S. et al. PheMap: a multi-resource knowledge base for high-throughput phenotyping within electronic health records. J. Am. Med. Inform. Assoc. 27, 1675–1687 (2020).

31. Carone, M., Petersen, M. & van der Laan M. Targeted minimum loss-based estimation of a causal effect using interval-censored time-to-event data. (2012).

32. Kreif, N. et al. Exploiting nonsystematic covariate monitoring to broaden the scope of evidence about the causal effects of adaptive treatment strategies. Biometrics 77, 329–342 (2021).

33. van der Laan, M. J. & Robins, J. M. Unified Methods for Censored Longitudinal Data and Causality. (Springer, New York, NY, 2011).

34. Butzin-Dozier, Z. et al. Causal inference via electronic health records in the National Clinical Cohort Collaborative: Challenges and solutions in Long COVID research. medRxiv 2025.06.06.25329168 (2025) doi:10.1101/2025.06.06.25329168.

35. Wickham, H. Ggplot2: Elegant Graphics for Data Analysis. (Springer, New York, NY, 2009).

36. OpenAI. https://openai.com/.

37. Claude. https://claude.ai/login?returnTo=%2F%3F.

38. Lee, S. Y. & Lee, K. Factors that influence an individual’s intention to adopt a wearable healthcare device: The case of a wearable fitness tracker. Technol. Forecast. Soc. Change 129, 154–163 (2018).

39. Pan, C.-C. et al. Sociodemographics and digital health literacy in using wearables for health promotion and disease prevention: Cross-sectional nationwide survey in Germany. J. Prev. 46, 371–391 (2025).

40. Dunn, J. et al. Defining the habitome: Phenotypes of routine and their relationship to health outcomes. Research Square (2025) doi:10.21203/rs.3.rs-5861743/v2.

41. Active ageing: A policy framework. Preprint at https://iris.who.int/bitstream/handle/10665/67215/WHO_NMH_NPH_02.8.pdf;jsessionid=05C9853E78DCB8F5182322FAF23C4798.

42. Moore, K. et al. Older adults’ experiences with using wearable devices: Qualitative systematic review and meta-synthesis. JMIR MHealth UHealth 9, e23832 (2021).

43. Li, J., Ma, Q., Chan, A. H. & Man, S. S. Health monitoring through wearable technologies for older adults: Smart wearables acceptance model. Appl. Ergon. 75, 162–169 (2019).

44. Lewis, Z. H., Pritting, L., Picazo, A.-L. & JeanMarie-Tucker, M. The utility of wearable fitness trackers and implications for increased engagement: An exploratory, mixed methods observational study. *Digit*. Health 6, 2055207619900059 (2020).

45. Chandrasekaran, R., Katthula, V. & Moustakas, E. Patterns of use and key predictors for the use of wearable health care devices by US adults: Insights from a national survey. J. Med. Internet Res. 22, e22443 (2020).

46. Holko, M. et al. Wearable fitness tracker use in federally qualified health center patients: strategies to improve the health of all of us using digital health devices. NPJ Digit. Med. 5, 53 (2022).

47. Cho, P. J. et al. Demographic imbalances resulting from the bring-your-own-device study design. JMIR MHealth UHealth 10, e29510 (2022).

48. Kim, E. H. et al. Association of demographic and socioeconomic indicators with the use of wearable devices among children. *JAMA Netw*. Open 6, e235681 (2023).

49. de Arriba-Pérez, F., Caeiro-Rodríguez, M. & Santos-Gago, J. M. Collection and processing of data from wrist wearable devices in heterogeneous and multiple-user scenarios. Sensors (Basel) 16, 1538 (2016).

50. Pitt, L. et al. Understanding the opportunities and challenges of wearable technology. in Creating Marketing Magic and Innovative Future Marketing Trends 139–141 (Springer International Publishing, Cham, 2017).

51. Balbim, G. M. et al. Using Fitbit as an mHealth intervention tool to promote physical activity: Potential challenges and solutions. JMIR MHealth UHealth 9, e25289 (2021).

52. Hacker, K. & Houry, D. Social needs and social determinants: The role of the centers for disease control and prevention and public health. Public Health Rep. 137, 1049–1052 (2022).

53. Nagata, J. M. et al. Sociodemographic correlates of contemporary screen time use among 9– and 10-year-old children. J. Pediatr. 240, 213–220.e2 (2022).

54. Riley, W. J. Health disparities: gaps in access, quality and affordability of medical care. Trans. Am. Clin. Climatol. Assoc. 123, 167–72; discussion 172–4 (2012).

55. Shcherbina, A. et al. Accuracy in wrist-worn, sensor-based measurements of heart rate and energy expenditure in a diverse cohort. J. Pers. Med. 7, 3 (2017).

56. Saneei, P. et al. Combined healthy lifestyle is inversely associated with psychological disorders among adults. PLoS One 11, e0146888 (2016).

57. Kafadar, D., Esen, A. D. & Arıca, S. Determining health-promoting behavior in smokers preparing to quit: a holistic and personalized approach. EPMA J. 10, 115–123 (2019).

58. Minen, M. T. & Stieglitz, E. J. Wearables for neurologic conditions: Considerations for our patients and research limitations: Considerations for our patients and research limitations. Neurol. Clin. Pract. 11, e537–e543 (2021).

59. Katapally, T. R. & Muhajarine, N. Towards uniform accelerometry analysis: a standardization methodology to minimize measurement bias due to systematic accelerometer wear-time variation. J. Sports Sci. Med. 13, 379–386 (2014).

60. Rosenbaum, P. R. & Rubin, D. B. The central role of the propensity score in observational studies for causal effects. Biometrika 70, 41–55 (1983).

61. Miller, G. D. et al. Effect of varying accelerometry criteria on physical activity: The look AHEAD study. Obesity (Silver Spring) 21, 32 (2012).

62. Bagley, S. C. & Altman, R. B. Computing disease incidence, prevalence and comorbidity from electronic medical records. J. Biomed. Inform. 63, 108–111 (2016).

63. Gianfrancesco, M. A. & Goldstein, N. D. A narrative review on the validity of electronic health record-based research in epidemiology. BMC Med. Res. Methodol. 21, 234 (2021).

64. Research Roundup: All of Us Participants’ Fitbit Data Drive New Research. https://allofus.nih.gov/article/announcement-research-roundup-all-us-participants-fitbit-data-drive-new-research.

65. Resources for Using Fitbit Data. User Support https://support.researchallofus.org/hc/en-us/articles/20281023493908-Resources-for-Using-Fitbit-Data.

