## Supplemental material for "Population differences in wearable device wear time: Rescuing data to address biases and advance health equity"

### Supplementary Methods and Results

#### Supplementary Methods

<u>Data pre-processing for covariates</u>

Age was determined by taking the mean age in years between the first date of recorded Fitbit data and last date of recorded Fitbit data in AoU (relative to the date of birth). We filtered on individuals less than 99 years old and categorized the age in years into increments of 10 years starting from *less than 20 years old*, *20-30 years old*, through *70-80 years old*, and *above 80 years old* to get a more granular assessment of its impact as a covariate. If age was left as a continuous variable, the model output would report the percentage of wear time associated with an increase in 1 year of age as opposed to being able to observe differences in wear time within different age groups.

SDoH measures of annual income, education, and health insurance were obtained using the *Basics* survey in AoU[27]. The questions analyzed included “What is your annual household income from all sources?” to assess income, “What is the highest grade or year of school you completed” to assess education, and “Are you covered by health insurance or some other kind of health care plan” to assess health insurance. For responses related to annual income, responses were discrete and ranged from *less than 10k* to *more than 200k*. For the survey question regarding education, the following responses were combined into a new response of *highest grade: grade 12, GED, or lower* to avoid any participant counts lower than 20 in compliance with AoU’s publication and dissemination policy: 1) *highest grade: five through eight*, 2) *highest grade: nine through eleven*, and 3) *highest grade: twelve or GED*. For the question about health insurance, we only included individuals who responded *yes* or *no* for analysis. Lastly, we ensured survey responses were at the time of Fitbit ownership by checking the date each participant responded to survey questions was between the first and last date of available Fitbit data.

Lifestyle factors related to tobacco and alcohol use were determined using the *Lifestyle* survey in AoU[28]. The specific questions analyzed were: Have *you smoked at least 100 cigarettes in your entire life?* for tobacco use and *In your entire life, have you had at least one 1 drink of any kind of alcohol, not counting small tastes of sips?*. Responses to both questions were binomial (i.e., yes or no). We filtered the data on 30 days of Fitbit data prior to the date of the survey to capture recent tobacco and alcohol use behavior and calculated the percentage of days and the daily hours of Fitbit wear time over that 30-day period. Linear regression models were used to analyze the percentage of days, and linear mixed-effects models for the daily hours, with a significance level of 0.05.

Preprocessing depressive, anxiety, and anhedonia symptoms

While the initial intentions of AoU conducting this survey were to assess underlying changes in participant experiences and health during the COVID-19 pandemic, we leveraged the questions related to mood, anxiety, and anhedonia for the purpose of investigating their relationship with Fitbit wear time in this analysis. Survey responses were used to ascertain whether an individual was experiencing either depressive, anxiety, and/or anhedonia symptoms. The survey questions about depressive, anxiety, and anhedonia symptoms were *Over the last 2 weeks, how often have you been bothered by the following problem: feeling down, depressed, or hopeless?*, *Over the last 2 weeks, how often have you been bothered by the following problem: feeling nervous, anxious, or on edge?*, and *Over the last 2 weeks, how often have you been bothered by the following problem: little interest in pleasure or doing things?*, respectively. Given that the surveys assessed mood, anxiety, and anhedonia symptoms over the previous two week period, we identified the survey date for each individual and calculated the percentage of days and hours per day they wore their Fitbit from 14 days before the survey date through the survey date. We included only individuals who took the survey after their first recorded Fitbit usage to ensure the availability of Fitbit data for analysis. We also filtered on individuals with at least 14 total days of Fitbit data to ensure each person had enough possible data for accurate analysis. Responses to mood and anxiety survey questions consisted of 1) *Not at all*, 2) *Several days*, *More than half the days*, and *Nearly every day*. First, we simplified the responses to a binary variable where those with a response of *Not at all* were considered not to have depressive, anxiety, or anhedonia symptoms (depending on the question), while those with any other response were considered to have symptoms. We then compared the percentage of days and hours per day individuals wore their Fitbit over the 14-

day period between individuals 1) with and without depressive symptoms, 2) with and without anxiety symptoms, and and 3) with and without anhedonia symptoms using linear regression models at a significance level of 0.05, adjusting for age, sex, and race/ethnicity (processed as described above). Next, recognizing the high rate of co-occurring anxiety and depressive symptoms[30,31], the data was reanalyzed combining binary responses from depressive and anxiety symptom survey questions to create groups of individuals with either 1) both depressive and anxiety symptoms, 2) only depressive symptoms, 3) only anxiety symptoms, or 4) neither depressive nor anxiety symptoms and compared using a linear regression model at a significance level of 0.05. Lastly, to directly compare the 1) percentage and 2) hours of daily Fitbit wear time amongst those with anxiety symptoms alone compared to depressive symptoms alone, we filtered on individuals with anxiety or depressive symptoms alone and ran additional models. All models described in this section were adjusted for age, sex, race/ethnicity, annual income, education level, and health insurance status.

<u>Preprocessing major depressive disorder and anxiety disorder diagnosis data</u>

Concepts including the name "remission" were excluded to restrict our analysis to those with current depressive symptoms. We filtered on the earliest record of a depression diagnosis for each individual. Individuals without depression were identified by taking those in the entire AoU cohort who were not part of the depression cohort. Fitbit data analyzed only included the 14 days prior to the date of diagnosis through the first 30 days after diagnosis for two reasons: 1) the diagnostic criteria for MDD requires that individuals experience symptoms for at least 14 days, so we assumed that individuals were experiencing depressive symptoms starting at 14 days prior to the index date, and 2) some individuals were given an antidepressant medication on the date of diagnosis which can take 4-6 weeks to take effect, prompting us to exclude data after 30 days past the index date to capture the window of when individuals were experiencing depressive symptoms, consistent with prior studies[19]. To establish a control group with an equivalent number of days of Fitbit data in the non-MDD cohort, we implemented established methods[19] and identified the median number of days from the first available Fitbit data to the MDD diagnosis for individuals in the MDD cohort. This median value was then used as the index date for individuals in the non-MDD cohort, along with the following 44 days of Fitbit data, ensuring a total of 45 days of data for both the MDD and non-MDD cohorts. To ensure accurate analysis, we filtered on individuals who had at least 45 total days of Fitbit data.

Individuals with anxiety disorder (AD) were identified using OMOP concept ID 442077, which includes all subtypes of anxiety disorders. Concepts including the terms *acute*, *child*, *adolescence*, and *physical and emotional exhaustive state* were excluded to better capture individuals who were experiencing anxiety disorders at the time of the analysis. Individuals without anxiety were identified in the same manner as the non-MDD cohort. We also followed a similar process in the survey data analysis to create groups of individuals with a diagnosis of either 1) both MDD and AD, 2) MDD alone, 3) AD alone, or 4) neither MDD nor AD. Individuals were only considered to be in group 1 (the MDD and AD cohort) if they were diagnosed with both conditions within three months of one another.

For analysis, the percentage of days and hours per day the Fitbit was worn was calculated as previously described using the 45 days of data for each cohort. Similarly, models comparing 1) individuals with versus without MDD and 2) individuals with versus without AD were run. We focused on the 14 days prior to the index date through 30 days after diagnosis, totaling 45 days of data and calculated the 1) percentage of days or 2) hours per day the Fitbit was worn. Models were then run with linear regression adjusted for age, sex, race/ethnicity, annual income, education level, and health insurance status at a significance level of 0.05 to compare the percentage of wear time between the four groups relative to those with neither MDD nor AD. To directly compare differences in wear time percentage between those with MDD only and AD only, we filtered our multinomial cohort on individuals diagnosed with MDD only or AD only and reran models adjusted for age, sex, race/ethnicity, annual income, education level, and health insurance status at a significance level of 0.05 with Bonferroni correction.

#### Supplementary Results

**Table S1: Phecode grouping of mental disorders included for analysis.**

| Phecode integer | Description | Count (%) |
|---|---|---|
| 290 | Delirium dementia and amnestic and other cognitive disorders | 124 (1.04%) |
| 291 | Other specified nonpsychotic and/or transient mental disorders | 275 (2.31%) |
| 292 | Neurological disorders | 604 (5.07%) |
| 293 | Symptoms involving head and neck | 425 (3.57%) |
| 295 | Schizophrenia and other psychotic disorders | 53 (0.45%) |
| 296 | Mood disorders | 1,764 (14.8%) |
| 297 | Suicidal ideation or attempt | 114 (0.96%) |
| 300 | Anxiety, phobic, and dissociative disorders | 2,041 (17.1%) |
| 301 | Personality disorders | 81 (0.68%) |
| 302 | Sexual and gender identity disorders | 190 (1.60%) |
| 303 | Psychogenic and somatoform disorders | 162 (1.36%) |
| 304 | Adjustment reaction | 675 (5.67%) |
| 305 | Eating disorders | 97 (0.82%) |
| 306 | Other mental disorders | 330 (2.77%) |
| 312 | Conduct disorders | 31 (0.26%) |
| 313 | Pervasive developmental disorders | 318 (2.67%) |
| 315 | Developmental delays and disorders | 50 (0.42%) |
| 316 | Substance addiction and disorders | 226 (1.90%) |
| 317 | Alcohol-related disorders | 205 (1.72%) |
| 318 | Tobacco use disorder | 420 (3.53%) |

Note: the percentage is calculated relative to the total cohort sample size (n=11,901).

**Table S2: Phecode grouping of all disorders included for analysis.**

| Group | Count (%) |
|---|---|
| None (healthy controls) | 5,688 (48.0%) |
| Blood/immune | 2,172 (18.3%) |
| Circulatory | 4,157 (35.1%) |
| Digestive | 3,926 (33.1%) |
| Dermatologic | 3,981 (33.6%) |
| Endocrine/metabolic | 4,534 (38.3%) |
| Genitourinary | 4,307 (36.3%) |
| Infectious diseases | 2,674 (22.6%) |
| Injuries and poisoning | 3,567 (30.1%) |
| Mental disorders | 3,335 (28.1%) |
| Musculoskeletal | 3,542 (29.9%) |
| Neurological | 4,841 (40.9%) |
| Neoplasms | 3,354 (28.3%) |
| Perinatal period | 3,649 (30.8%) |
| Pregnancy complications | 630 (5.3%) |
| Respiratory | 4,396 (37.1%) |
| Symptoms | 4,094 (34.6%) |

**Figure S1: Differences in the percentage of days individuals wear their Fitbit by lifestyle factors.**

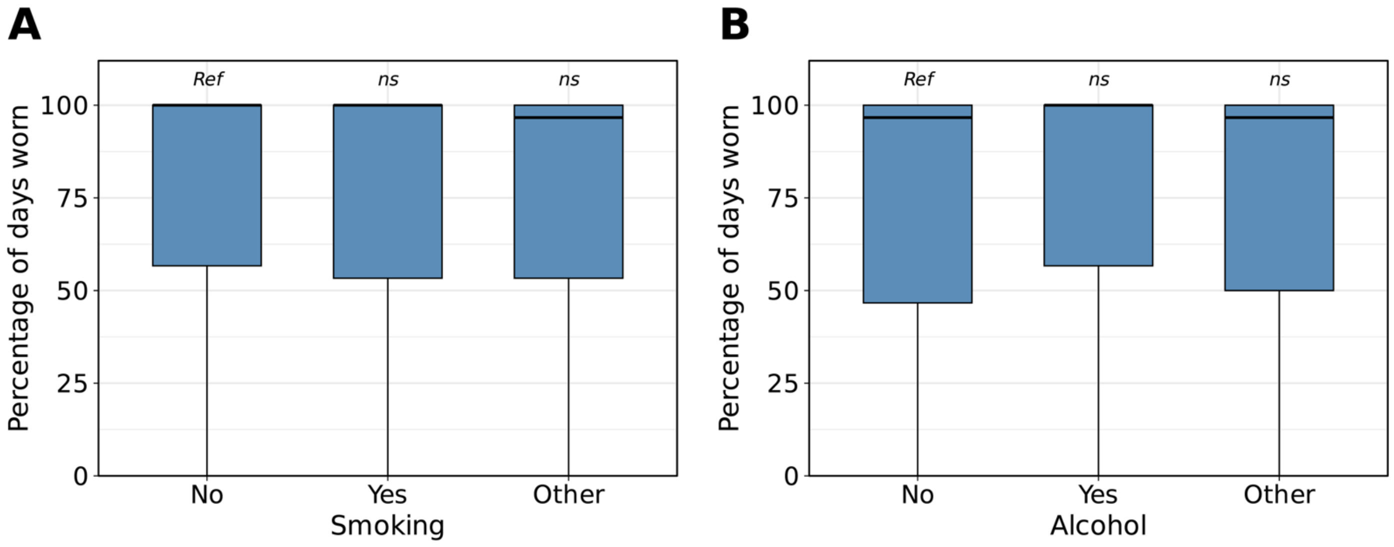


*ns*=not significant, *=*P*<0.05, **=*P*<0.01, ***=*P*<0.001

*The percentage of days individuals in the AoU cohort wore their Fitbit by smoking status (A) and alcohol status (B). Data in each group were compared using linear regression and are expressed as mean and 95% confidence intervals. Statistical tests were run relative to a reference group represented as "Ref". There was no significant difference in the percentage of days individuals wore their device by smoking or alcohol status.*

**Figure S2: Differences in the hours per day individuals wear their Fitbit by lifestyle factors.**

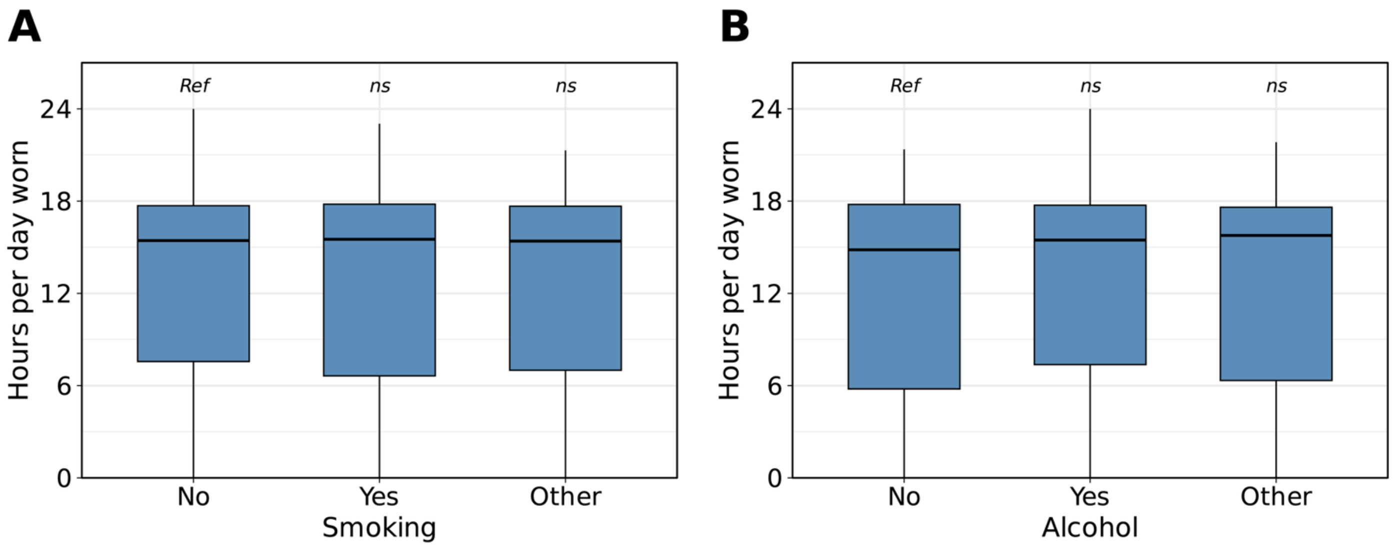


*ns*=not significant, *=*P*<0.05, **=*P*<0.01, ***=*P*<0.001

*The hours per day individuals in the AoU cohort wore their Fitbit by smoking status (A) and alcohol status (B). Data in each group were compared using linear regression and are expressed as mean and 95% confidence intervals. Statistical tests were run relative to a reference group represented as "Ref". There was no significant difference in the hours per day individuals wore their device by smoking or alcohol status.*

**Figure S3: Association between mental disorder categories and Fitbit wear time before and after diagnosis.**

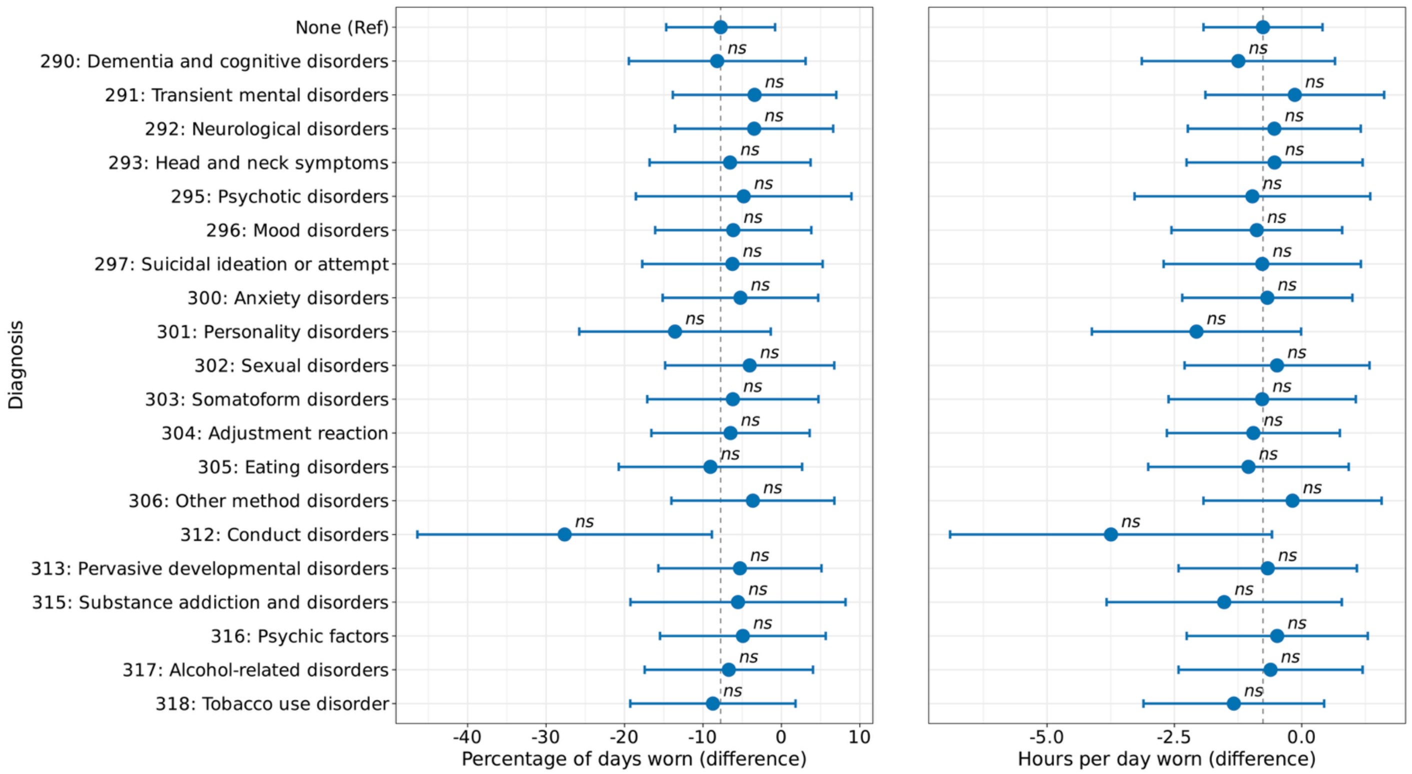


*ns*=not significant, *=*P*<0.05, **=*P*<0.01, ***=*P*<0.001 after Bonferroni correction

*The difference in the percentage of days (A) and hours per day (B) individuals in the AoU cohort wore their Fitbit by mental health disorders before and after diagnosis. We used linear mixed-effects models with diagnosis-by-period interactions and random intercepts for person ID to test whether changes in device wear time (percentage of days and hours per day) differed between diagnostic groups and controls. All models adjusted for age, sex, race/ethnicity, annual income, education level, and health insurance status. Data are expressed as mean and 95% confidence intervals. For each mental health disorder, the interaction model compared post-diagnosis wear time to the index date among controls without a mental health disorder (reference period, indicated by the dashed line). Fitbit device wear time did not differ between mental disorder groups and controls post-diagnosis.*

**Figure S4: Association between chronic disease categories and Fitbit wear time before and after diagnosis.**

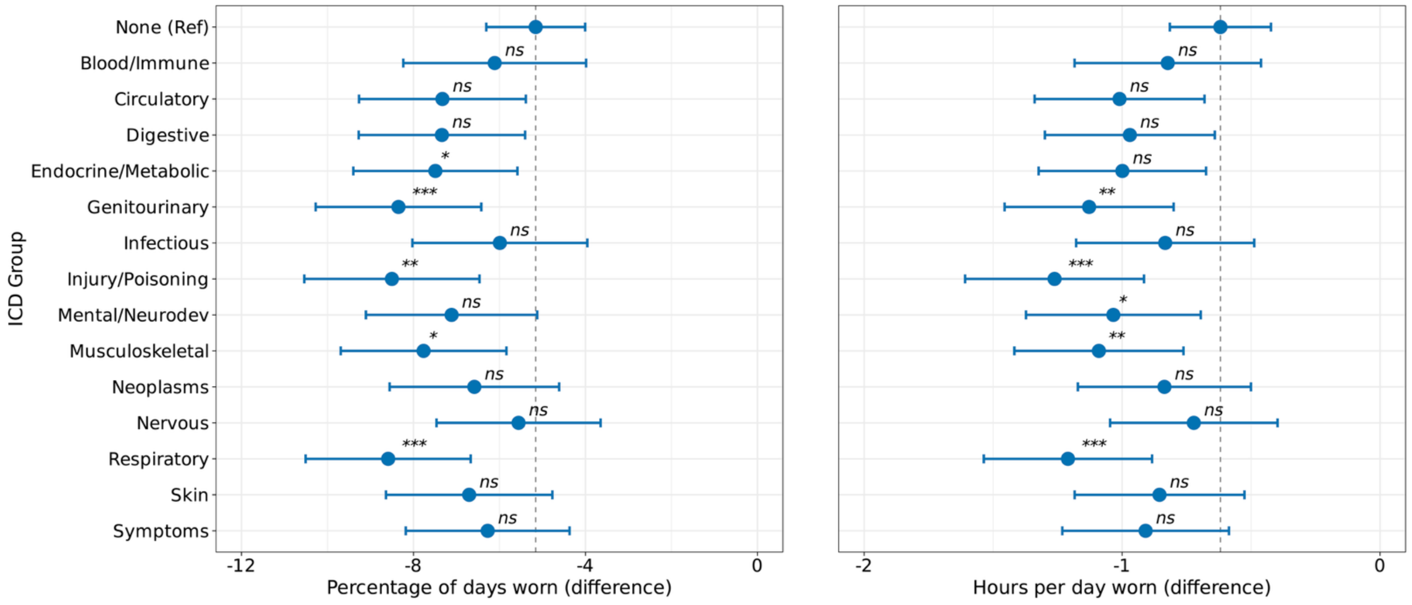


*ns*=not significant, *=*P*<0.05, **=*P*<0.01, ***=*P*<0.001

*The difference in the percentage of days (A) and hours per day (B) individuals in the AoU cohort wore their Fitbit by disease groups before and after diagnosis. We used linear mixed-effects models with diagnosis-by-period interactions and random intercepts for person ID to test whether changes in device wear time (percentage of days and hours per day) differed between diagnostic groups and controls. All models adjusted for age, sex, race/ethnicity, annual income, education level, and health insurance status. Data are expressed as mean and 95% confidence intervals. For each chronic disease group, the interaction model compared post-diagnosis wear time to the index date among controls without any diagnosed chronic disease (reference period, indicated by the dashed line). Endocrine/metabolic, genitourinary, injury/poisoning, musculoskeletal, and respiratory diagnoses were associated with greater declines after the index date in device wear compared to controls.*
